# Transdiagnostic Domain-specific Cognitive Impairment in Inter-episode Mood Disorders and its Relationship with Rest-Activity Phenotypes

**DOI:** 10.64898/2026.08.16.26360541

**Authors:** Filippo Corponi, Michail Kalfas, Matteo Reami, Giuseppe Fanelli, Paolo Ossola, Sameer Jauhar, Allan H Young

## Abstract

**Introduction:** Cognitive impairment and disturbed rest-activity patterns often persist between episodes of major depressive disorder (MDD) and bipolar disorder (BD). Whether these deficits are disorder-specific or transdiagnostic remains unclear, as does the existence of a link between rest-activity phenotypes and cognitive performance.

**Methods:** Using the *All of Us* Research Program, we derived normative deviation scores across four cognitive domains (sustained attention, inhibitory control, reward-based impulsivity, social cognition) from non-clinical controls (NCC), then compared deviations in MDD and BD. MDD was adequately powered to regress deviation scores on four 90-day Fitbit-derived phenotypes (step count, sleep duration, wakefulness after sleep onset, sleep timing variability); the same analysis was run on NCC as a sensitivity check.

**Results:** Samples were substantially larger than prior works (MDD 5,087–6,536; BD 545–739; NCC 40,589–51,491). Relative to NCC, MDD and BD exhibited worse sustained attention (Δ_Glass_ = 0.081 vs. 0.187) and higher impulsivity (Δ_Glass_ = 0.092 vs. 0.240), with deficits more pronounced in BD. No wearable phenotype was significantly associated with cognitive performance in MDD (*R*^2^ *<* 0.01); NCC associations, though significant, were of negligible magnitude (*R*^2^ *≤* 1.2%).

**Discussion:** Inter-episode cognitive impairment was domain-selective rather than global, with a gradient BD *>* MDD. Despite adequate power, wearable rest-activity phenotypes were not associated with cognition in MDD. Whether this extends to BD, where deficits were largest, could not be tested due to limited power. Community-dwelling samples likely underestimate impairment relative to clinical cohorts.

## 1 Introduction

Major depressive disorder (MDD) and bipolar disorder (BD) are two of the most prevalent and debilitating psychiatric conditions [1, 2]. Both have historically been conceptualised as mood (or affective) disorders, as pathological mood shifts are amongst the most prominent features of their clinical presentation. Both disorders follow an episodic, relapsing-remitting course [3, 4], with acute episodes giving way to more stable inter-episode periods known as euthymia. Major depressive episodes occur in both MDD and BD, whereas (hypo)manic episodes, marked by elevated or irritable mood, are specific to BD.

Historically, clinical management has focused primarily on resolving acute mood symptoms and preventing relapse. However, a growing body of evidence shows that syndromal remission does not equate to full functional recovery: up to 60% of individuals with MDD and BD continue to experience significant functional impairment in social and occupational domains despite the resolution of mood symptoms [5–7]. This gap between symptomatic remission and functional recovery has shifted research attention toward mechanisms that persist beyond acute episodes and may contribute to incomplete functional recovery or vulnerability to recurrence.

In euthymic BD, an estimated 37-64% of individuals report cognitive difficulties [8], with cognitive impairment also commonly reported in remitted MDD [9]. These deficits may persist beyond the resolution of mood symptoms [10, 11] and are associated with poorer psychosocial functioning [12, 13]. Particularly in affective disorders, cognition can be conceptualised in terms of partially overlapping cold, hot, and social domains [14, 15]. Cold cognition encompasses relatively affect-neutral information processing; hot cognition involves affectively or motivationally salient information; and social cognition concerns the perception and interpretation of socially relevant information, drawing on both cold and hot processes [16]. Compared with non-clinical controls, individuals with euthymic BD [17, 18] and remitted MDD [19–21] show impairments across several domains of cold cognition, including executive functioning, attention, verbal memory, and working memory. Similarly, altered processing of emotionally laden information has been observed during euthymia in BD [22] and remission in MDD [23], with the latter more consistently characterised by preferential processing of negative over positive information. Finally, social-cognitive alterations may also persist beyond acute episodes. In euthymic BD, evidence indicates impairments in mentalising and facial-emotion recognition [24–26], whereas the more limited and heterogeneous literature on remitted MDD suggests persistent alterations in mentalising and the processing of emotional social cues [27, 28]. Direct comparative studies have generally found similar performance across most cold-cognitive domains, although preliminary evidence suggests greater verbal-memory impairment in BD [29, 30].

Importantly, these impairments are clinically relevant even during euthymia, when syndromal mood symptoms have resolved. Cold-cognitive deficits may be reflected in persistent and perceived difficulties with concentration, planning, and memory [31]. Alterations in hot cognition may parallel residual affective tendencies: reduced reward sensitivity may contribute to anhedonia and low motivation [32]; preferential processing of negative information may confer vulnerability to depressive recurrence [33]; and heightened reward sensitivity may be associated with subsyndromal activation in BD [34]. Finally, social-cognitive difficulties may manifest as difficulties in adjusting behaviour to social feedback, potentially contributing to withdrawal, and impaired social functioning [35]. These abnormalities should not necessarily be regarded as residual symptoms themselves; rather, they may represent trait-like vulnerabilities linking residual symptomatology to functional impairment and future relapse [36]. Accordingly, comparing non-clinical controls, remitted MDD, and euthymic BD across cold, hot, and social cognition in a large, well-characterised sample may clarify whether persistent cognitive alterations are shared across affective disorders or distinguish MDD from BD, while overcoming the limited sample sizes and methodological heterogeneity of previous meta-analysed studies [36].

Building on Kraepelin’s classical description of BD, psychomotor activation has recently gained prominence as a central feature in the characterisation of affective disorders [37, 38]. More specifically, alterations in psychomotor activity and circadian rhythms have been hypothesised to precede cognitive and affective symptoms during the emergence of major mood episodes [39]. Sleep-wake and activity disruption may represent one mechanism associated with persistent cognitive impairment, although their contribution independently of illness history and residual symptoms remains unclear.

Few studies have directly examined the relationship between sleep and cognition in mood disorders. A systematic review of this literature [40] found that studies using subjective sleep measures generally reported associations between sleep disturbance and cognitive impairment in both MDD and BD. By contrast, evidence based on objective sleep measures was sparse and inconsistent. In remitted MDD, only one study combined polysomnography with cognitive assessment in a sample of 57 participants, finding independent associations of depression history and poorer sleep with psychomotor slowing, but no evidence that sleep moderated depression-related impairments in executive control [41]. Other objective studies in MDD have produced similarly mixed findings. A twelve-month actigraphy study found that within-person rest-activity fluctuations tracked both depressive severity and attention across depressive and remitted periods [42], whereas a larger consumer-wearable study found no association between physiologically measured sleep and neurocognitive performance despite a correlation with self-reported sleep disturbance [43]. Overall, the evidence remains fragmented, based largely on small samples and subjective sleep reports; studies using objective measures have generally examined limited cognitive domains over short recording periods.

Beyond the sleep-wake cycle, physical activity may also be relevant, given its associations with psychomotor functioning, circadian rhythms, and cognition; although systematic reviews have characterised actigraphy-derived rest-activity abnormalities in euthymic BD and remitted MDD [44–46] no review has specifically synthesised the relationship between objectively measured habitual physical activity and cognitive functioning at the remission of an acute affective episode.

### Aims

Using data from the All of Us Research Program [47], the present study addressed two aims. First, we compared non-clinical controls, individuals with remitted MDD, and individuals with euthymic BD across cold, hot, and social cognition to determine whether persistent cognitive alterations are shared across affective disorders or distinguish MDD from BD. Second, within remitted MDD, we explored whether performance across these cognitive domains was associated with wearable-derived sleep estimates derived from 90 days of Fitbit recordings preceding the cognitive test date. Together, these analyses examined whether behavioural and sleep-wake rhythms contribute to heterogeneity in cognitive functioning after symptomatic remission.

## 2 Methods

### 2.1 Cohort

The *All of Us* Research Program is a US-based longitudinal cohort. A description of this cohort was given elsewhere [47]. This analysis used data from Curated Data Repository version 9 (CDRv9 - C2025Q4R6). Physical activity and sleep were monitored using Fitbit^TM^ devices under either the “Bring Your Own Device” model (participants linking pre-existing accounts) or the 2021 Wearables Enhancing All of Us Research (WEAR) study, which provided devices to underrepresented populations.

### 2.2 Cognitive Tasks: the *Exploring the Mind* Battery

Cognitive function was assessed using the *Exploring the Mind* (EtM) battery [48], a suite of four computerised tasks administered remotely and unsupervised through the *All of Us* participant portal. Developed jointly by the *All of Us* Research Program and the National Institute of Mental Health (NIMH), the battery probes transdiagnostic functional domains specified by the Research Domain Criteria (RDoC) framework [49, 50]. All tasks had been validated for large-scale remote administration via TestMyBrain, with performance comparable to laboratory-based administration [51, 52]. Tasks were available from September 2023 and could be completed in any order on a personal device.

The **Gradual-Onset Continuous Performance Task** (*City or Mountain*, **GradCPT**) measured response inhibition in attentional control, that is the ability to suppress or withhold a prepotent, automatic, or inappropriate motor response. Participants viewed a four-minute stream of 300 greyscale images transitioning every 800 ms between city scenes (89.3% of trials; go stimuli) and mountain scenes (10.7%; no-go stimuli), responding to cities while withholding responses to mountains. The primary outcome was *d^→^* (dprime), a signal-detection index of discrimination sensitivity, with higher values indicating better performance. This index is computed as the difference between the hits, the correct presses, and the false alarms, the incorrect presses, both z-scored within individuals.

The **Flanker Attention Task** (*Left or Right*, **Flanker**) measured attentional inhibition in attentional control, that is the ability to allocate and maintain attention on goal-relevant information while minimising distraction from irrelevant stimuli. Across 96 scored trials, participants indicated the direction of a central target arrow flanked on each side by two arrows pointing either in the same direction (congruent trials) or on the opposite direction (incongruent trials).

The primary outcome was rate-correct-score interference (RCS_int_), the difference in accuracy between congruent and incongruent trials; higher values indicate greater flanker interference and worse cognitive control. To account for inter-individual differences, both accuracies are divided by the subject’s median reaction time in seconds.

The **Delay Discounting Task** (*Now or Later*, **DD**) measured temporal discounting in reward-based decision making, that is, the tendency of people to perceive a reward as less valuable the further in the future it occurs. Using an adaptive staircase, participants chose between a smaller immediate reward and $1,000 after one of four hypothetical delays (two weeks, one month, one year, ten years) across 24 trials. The subjective value of the delayed reward was assumed to decline as a hyperbolic function of delay, with the discount rate (*k*) quantifying how rapidly future rewards lost subjective value. The primary outcome was ln *k*, the natural logarithm of the mean discount rate across the four delay periods; higher values indicate steeper temporal discounting, reflecting a stronger preference for smaller immediate rewards over larger delayed rewards.

The **Multiracial Facial Emotion Recognition Task** (*Guess the Emotion*, **EmoRecog**) measured facial emotion labelling in social cognition, that is, the ability to accurately identify and interpret emotional facial expressions. Looking at 48 photographs of demographically diverse actors, participants identified which of four emotions was expressed: happiness, sadness, anger, fear. We computed the average accuracy across the four emotions, i.e. EmoRec = (*A*_happy_ + *A*_sad_ + *A*_fearful_ + *A*_angry_)*/*4, where *A_e_* denotes proportion correct for emotion *e*. Higher scores indicate greater accuracy in labelling emotional expressions.

Within the cold–hot–social framework outlined above, the GradCPT and Flanker tasks indexed relatively affect-neutral attentional control (cold cognition), the Delay Discounting Task indexed reward-based decision-making (hot cognition), and the Facial Emotion Recognition Task indexed social cognition. Each of the cognitive tasks just described was modelled independently but underwent the same analytical pipeline outlined below.

### 2.3 Inclusion & Exclusion Criteria

#### 2.3.1 Participants

Psychiatric diagnoses were ascertained from Electronic Health Records (EHR) using standardised concept identifiers from the Observational Medical Outcomes Partnership (OMOP) Common Data Model, which harmonises SNOMED CT, ICD-9-CM, and ICD-10-CM vocabularies [53] (Table S2).

Participants were eligible for the MDD cohort if they had at least one pre-test EHR record of major depressive disorder (a threshold consistent with prior studies using *All of Us* Research Program [54–56]) and no history of bipolar disorder. Participants with at least one pre-test record of bipolar disorder were assigned to the BD cohort; those with records of both MDD and BD were assigned exclusively to BD.

Both clinical cohorts excluded individuals with EHR records of schizophrenia or schizoaffective disorder, attention deficit hyperactivity disorder, autism spectrum disorder, cognitive impairment, or drug dependence (excluding nicotine or tobacco dependence), as these conditions have distinct cognitive profiles that could confound mood-disorder-specific trait effects.

A Non-Clinical Controls (NCC) cohort provided a normative reference for task performance. NCC were required to have no EHR record of any mental health disorder. As in the MDD cohort, nicotine and tobacco dependence were permitted, while dementia and cognitive impairment were excluded.

#### 2.3.2 EtM Battery, Fitbit Recordings, and Inter-episode Phase Definition

Where a participant completed a given EtM task more than once (permitted after a mandatory 30-day inter-session interval), only the first administration was retained to avoid learning effects. EtM sessions were subjected to platform-provided quality-control flags [48, 57] (Table S1); sessions with at least one flag equal to one were excluded.

To ensure that cognitive and wearable phenotypes reflected trait-level inter-episode (euthymic) characteristics rather than (peri)episode states, EHR records were used in the absence of structured psychiatric state ratings in the *All of Us* Research Program. MDD and BD participants were excluded from the cognitive analyses if their EtM session occurred within 90 days before or after any EHR-recorded mood episode. The forward exclusion window was included because subsyndromal or prodromal states preceding a diagnosable episode may impair cognition before diagnostic criteria are met, introducing episode-state variance into a trait-level analysis.

For wearable analyses, the feature extraction window comprised the 90 calendar days immediately preceding the first EtM task administration. Participants were excluded if this 90-day window began within 90 days of an EHR-recorded mood episode, thereby excluding recent or ongoing episodes from the wearable phenotype.

### 2.4 Fitbit Data Processing

Days and nights within each participant’s 90-day window were quality-controlled before feature extraction, similarly to previous works [58–61]. A day was considered valid if heart rate (HR) data were recorded for at least 10 hours and the total step count fell between 250 and 45,000 steps. A night was considered valid if the main sleep episode had an onset time between 18:00 and 08:00 and a total sleep time between 120 and 720 minutes. Participants with fewer than 30 valid days or nights within the recording window were excluded from all wearable analyses.

Four phenotypes were derived. **Average Daily Step Count (Steps**, volume of physical activity), **Average Wakefulness After Sleep Onset** (**WASO**, the total minutes spent awake after initially falling asleep), and **Average Total Sleep Time** (**TST**, the total minutes spent asleep) were computed as arithmetic means across valid days and valid nights, respectively. **Sleep Timing Variability (STV)** was derived from the successive differences in sleep onset and offset times across consecutive valid nights. To avoid circular wraparound for sleep episodes spanning midnight, timing values were converted to noon-anchored decimal hours similarly to [62]. Under this convention, the recording day begins at 12:00 (noon), so that a sleep onset at 23:00 is represented as 23.0 and one at 01:00 the following calendar morning as 25.0. Formally, letting *H* and *M* denote the clock hour and minute of a given timestamp:

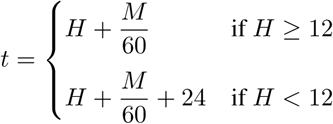

Successive differences in *t* were computed separately for onset (MASD_onset_) and offset (MASD_offset_) across strictly consecutive calendar nights; non-consecutive pairs were skipped to avoid artificial inflation of variability. STV was computed as the mean absolute successive difference (MASD), averaged across both channels:

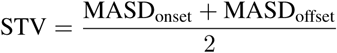

Higher STV values indicate greater sleep timing irregularity. STV was only computed for participants with at least 15 strictly consecutive valid nights within the recording window.

### 2.5 Statistical Analysis

The analyses presented below were conducted independently for each of the four tasks in the *EtM* battery. We do not investigate correlations between tasks (e.g., whether a participant’s deviation on GradCPT relates to their deviation on Flanker) but we treat each task as a separate measure of a specific cognitive dimension.

#### 2.5.1 Cognitive Performance Relative to Non-Clinical Controls in MDD and BD

Raw EtM task scores of MDD and BD participants were expressed as a normative deviation score *D*, representing the number of residual standard deviations by which an individual’s performance differed from NCC matched on age, sex, education, and device type (touchscreen or not) of test administration (Figure 1). Specifically, NCC raw scores were regressed on age (modelled with a cubic spline to allow a non-linear age-cognition relationship), sex, highest educational attainment (eight-level ordinal variable, from 0 = never attended school to 7 = advanced degree), and whether the EtM session was completed on a touchscreen device (to account for systematic differences in input latency and response speeds across hardware types [63]). A normative deviation score (*D*) was then derived for each clinical participant:

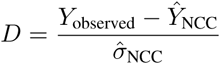

where *Y*_observed_ is the participant’s raw task score, *Ŷ*_NCC_ is the score predicted by the NCC regression for an individual the same age, sex, educational level, and device type, and 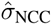 is the residual standard deviation of the NCC model. By construction, *D* is expressed in NCC-standardised units: it represents how many standard deviations an individual’s performance falls above or below their demographically matched healthy peers. Consequently, a *D* value of zero indicates performance indistinguishable from demographically matched NCC, while positive and negative values indicate scores above and below this expectation, respectively. Because higher raw scores reflect better performance for GradCPT and EmoRecog but worse performance (greater interference, steeper discounting) for Flanker and DD, the clinical interpretation of a positive or negative *D* (as better or worse relative to NCC) differs by task.

**Figure 1:**
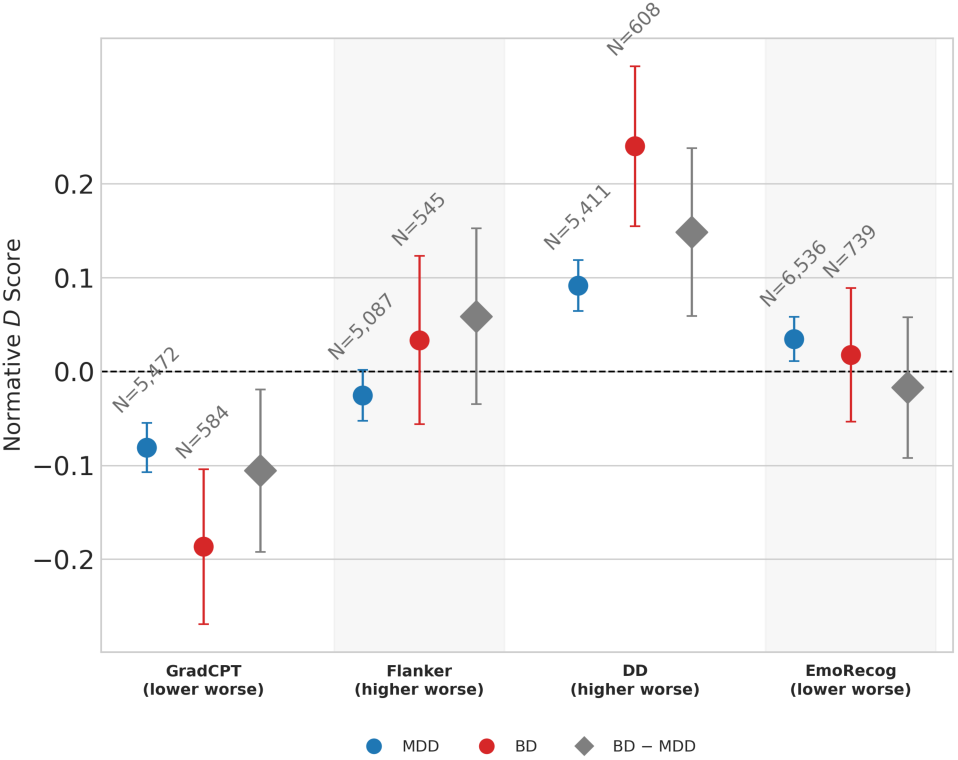
Normative deviation (*D*) across *EtM* battery tasks in MDD and BD. Point estimate of the average *D* and its 95% CI are displayed for Major Depressive Disorder (MDD, blue), Bipolar Disorder (BD, red), and the between-group contrast (BD − MDD, grey diamonds). Vertical grey shading highlights task blocks. Direction of impairment (*D* sign indicating worse performance) is indicated below each task label.

To assess how both clinical groups performed relative to NCC and to each other, we fitted a linear regression model containing only indicators for group membership (MDD and BD). This directly yielded the mean demographic-adjusted deviation for each diagnosis. We then used a linear contrast test on these coefficients to evaluate the direct difference between the two conditions (BD − MDD). To safeguard against unequal group sizes and unequal variances, all models utilised robust (HC3) standard errors.

#### 2.5.2 Wearable Phenotypes and Cognitive Performance in MDD

To investigate how habitual rest-activity phenotypes correlate with cognitive performance, we examined the relationship between cognitive deviations (*D*) and the four wearable metrics described above (Section 2.4: Steps, TST, WASO, and STV). Because only a fraction of EtM participants possessed (valid) Fitbit data, the sample available for this analysis is a subset of the larger cohort analysed in Section 2.5.1. In this context, individuals with BD were excluded entirely from this phase of the study because the BD wearable-eligible sample was severely underpowered to detect plausible effect sizes, as confirmed by the study of Minimum Detectable Effect Sizes for the realised sample size (Section 2.6).

To ensure the behavioural markers were comparable across participants in the MDD cohort, raw rest-activity phenotypes within this population were first residualised on age, sex, body mass index, and the Charlson Comorbidity Index [64] (CCI, accounting for non-mental comorbidities, e.g., cardiovascular or musculoskeletal co-occurring conditions), and then scaled to unit variance. Seasonal confounding, arising because the 90-day tracking window could fall across different times of the year, was addressed in the same regression by including the photoperiod at each participant’s recording midpoint. This photoperiod was computed via the Spencer [65] and Almorox [66] methods as implemented in the `chillR` package [67], with regional latitude derived from the first three digits of self-reported ZIP codes via `pgeocode` [68]. Consequently, the resulting standardised phenotype residuals capture individual variations in rest-activity phenotypes independent of these demographic, health, and seasonal factors.

Cognitive deviation (*D*) was then regressed on these standardised phenotype residuals, separately for each phenotype and task. The magnitude of each association was expressed as the Pearson correlation coefficient (*r*) between *D* and the residualised phenotype.

#### 2.5.3 Multiple Testing

The primary analysis comprised 28 distinct statistical tests. This total included 12 tests from Section 2.5.1 (evaluating the MDD group, the BD group, and the BD MDD contrast across the 4 cognitive tasks) and 16 tests from section 2.5.2 (evaluating the associations between the 4 rest-activity phenotypes and the 4 tasks). All *p*-values across these 28 tests were corrected jointly using the Benjamini–Hochberg procedure to control the false discovery rate (FDR) at 5%.

### 2.6 Minimum Detectable Effect Sizes

We computed the minimum detectable effect size (MDES) corresponding to 80% power for the realised sample size of each planned statistical test. Crucially, these calculations depend only on the sample size, significance threshold, and test specification, and not on the observed effect estimates [69].

Because exact statistical power cannot be derived analytically under the Benjamini-Hochberg false discovery rate (FDR) procedure, we bounded the MDES using two reference significance thresholds. The conservative bound assumes a Bonferroni correction across the 28 primary tests (*ε*_Bonf_ = 0.05*/*28 = 0.0018), whereas the optimistic bound assumes the nominal significance level (*ε*_nom_ = 0.05). The effective rejection threshold under the Benjamini-Hochberg procedure lies between these extremes, as do the corresponding MDES values.

### 2.7 Sensitivity Analysis

Three sensitivity analyses were performed to evaluate the robustness and representativeness of our findings, with all exploratory results evaluated at nominal *p*-values only (*p <* 0.05).

First, to confirm that the MDD subgroup with usable Fitbit data was representative of the broader EtM-tested MDD cohort, we conducted a cohort-equivalence check. We systematically compared the distribution of demographic (age, sex, education, test administration mode, BMI, physical comorbidity burden) and cognitive (baseline cognitive deviation *D*) features across the "Fitbit-eligible" against the "Fitbit-ineligible" MDD subsets.

Second, we re-estimated the phenotype–cognition regressions within the healthy control sample to serve as a non-clinical comparison arm, utilising the identical residualisation and regression pipeline described in Section 2.5.2.

Third, since both insufficient and excessive sleep can have detrimental health effects [70], we tested for non-linear relationships. The TST regression model, in both the MDD and NCC samples, was re-estimated with an added quadratic term to check for U-shaped or inverted U-shaped associations, particularly focusing on TST.

## 3 Results

### 3.1 Cognitive Performance Deviations in MDD and BD

The NCC reference population, used to compute normative *D* scores while controlling for age, sex, education level, and touchscreen use, comprised 43,515 individuals for GradCPT, 40,589 for Flanker, 42,734 for Delay Discounting, and 51,491 for EmoRecog. Demographic characteristics of the NCC sample are reported in Supplementary Material (Table S4). Sample sizes differ across *EtM* tasks because participants were free to complete any combination of tasks, not necessarily on the same day. Each task was modelled independently, in parallel, applying an identical analysis pipeline to each.

Sample retention through the inclusion and exclusion criteria described above is shown across all three groups (NCC, MDD, BD) in Supplementary Material (Table S3). Realised MDD and BD sample sizes were, respectively, 5,472 and 584 for GradCPT, 5,087 and 545 for Flanker, 5,411 and 608 for Delay Discounting, and 6,536 and 739 for EmoRecog. Clinical and demographic characteristics of the MDD and BD samples are shown in Table 1. The distribution of age, sex, education, and administration interface (touchscreen vs. standard) in NCC spans the ranges observed in MDD and BD, supporting the use of the NCC-derived normative model to compute demographically matched *D* scores for clinical participants without requiring substantial extrapolation beyond the range of the data on which the model was fit.

**Table 1:**
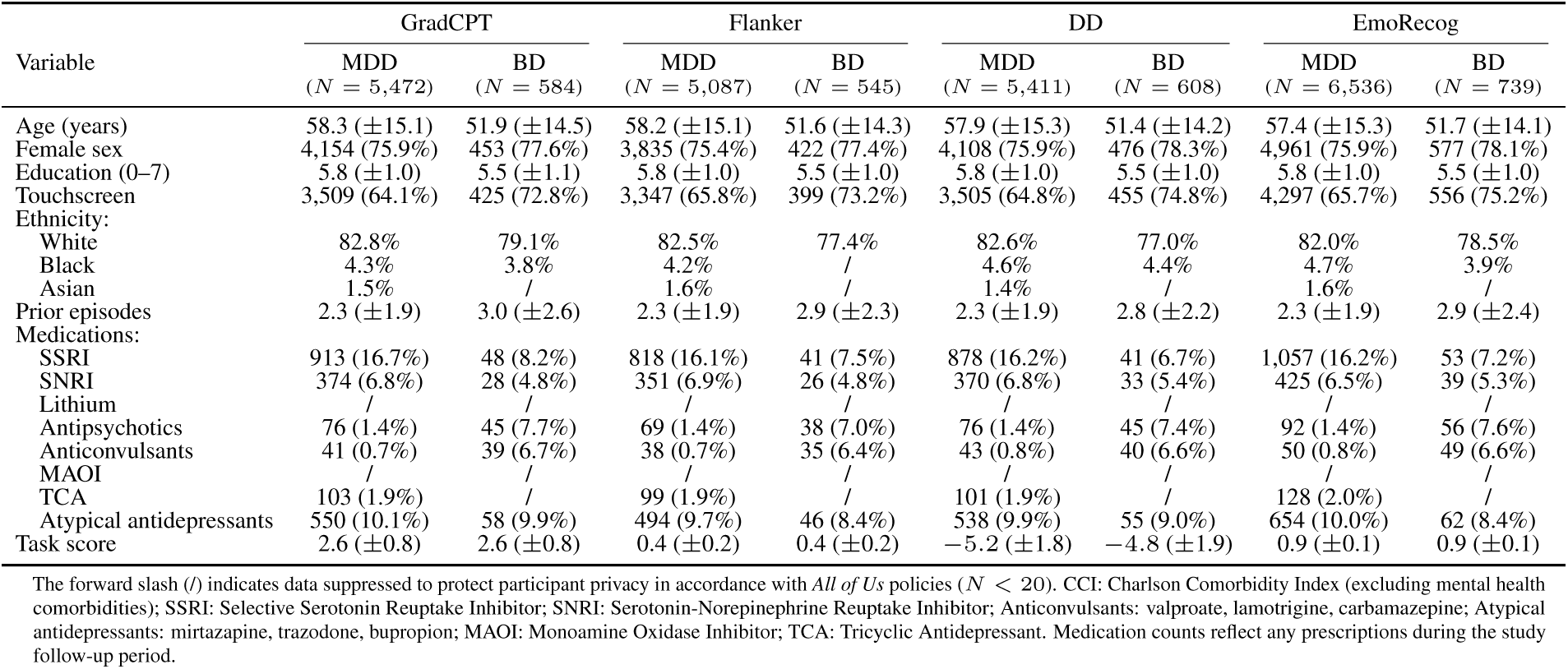
Analysis of Cognitive Performance Deviations: Clinical-demographics of MDD and BD cohorts across *EtM* tasks. Prior episodes: Derived from EHR data using a heuristic that consolidated diagnostic entries within 180 days into a single episode.

| Variable | GradCPT |  | Flanker |  | DD |  | EmoRecog |  |
| --- | --- | --- | --- | --- | --- | --- | --- | --- |
| | MDD<br>($N = 5,472$) | BD<br>($N = 584$) | MDD<br>($N = 5,087$) | BD<br>($N = 545$) | MDD<br>($N = 5,411$) | BD<br>($N = 608$) | MDD<br>($N = 6,536$) | BD<br>($N = 739$) |
| Age (years) | 58.3 ($\pm 15.1$) | 51.9 ($\pm 14.5$) | 58.2 ($\pm 15.1$) | 51.6 ($\pm 14.3$) | 57.9 ($\pm 15.3$) | 51.4 ($\pm 14.2$) | 57.4 ($\pm 15.3$) | 51.7 ($\pm 14.1$) |
| Female sex | 4,154 (75.9%) | 453 (77.6%) | 3,835 (75.4%) | 422 (77.4%) | 4,108 (75.9%) | 476 (78.3%) | 4,961 (75.9%) | 577 (78.1%) |
| Education (0–7) | 5.8 ($\pm 1.0$) | 5.5 ($\pm 1.1$) | 5.8 ($\pm 1.0$) | 5.5 ($\pm 1.0$) | 5.8 ($\pm 1.0$) | 5.5 ($\pm 1.0$) | 5.8 ($\pm 1.0$) | 5.5 ($\pm 1.0$) |
| Touchscreen | 3,509 (64.1%) | 425 (72.8%) | 3,347 (65.8%) | 399 (73.2%) | 3,505 (64.8%) | 455 (74.8%) | 4,297 (65.7%) | 556 (75.2%) |
| Ethnicity: |  |  |  |  |  |  |  |  |
| White | 82.8% | 79.1% | 82.5% | 77.4% | 82.6% | 77.0% | 82.0% | 78.5% |
| Black | 4.3% | 3.8% | 4.2% | / | 4.6% | 4.4% | 4.7% | 3.9% |
| Asian | 1.5% | / | 1.6% | / | 1.4% | / | 1.6% | / |
| Prior episodes | 2.3 ($\pm 1.9$) | 3.0 ($\pm 2.6$) | 2.3 ($\pm 1.9$) | 2.9 ($\pm 2.3$) | 2.3 ($\pm 1.9$) | 2.8 ($\pm 2.2$) | 2.3 ($\pm 1.9$) | 2.9 ($\pm 2.4$) |
| Medications: |  |  |  |  |  |  |  |  |
| SSRI | 913 (16.7%) | 48 (8.2%) | 818 (16.1%) | 41 (7.5%) | 878 (16.2%) | 41 (6.7%) | 1,057 (16.2%) | 53 (7.2%) |
| SNRI | 374 (6.8%) | 28 (4.8%) | 351 (6.9%) | 26 (4.8%) | 370 (6.8%) | 33 (5.4%) | 425 (6.5%) | 39 (5.3%) |
| Lithium | / | / | / | / | / | / | / | / |
| Antipsychotics | 76 (1.4%) | 45 (7.7%) | 69 (1.4%) | 38 (7.0%) | 76 (1.4%) | 45 (7.4%) | 92 (1.4%) | 56 (7.6%) |
| Anticonvulsants | 41 (0.7%) | 39 (6.7%) | 38 (0.7%) | 35 (6.4%) | 43 (0.8%) | 40 (6.6%) | 50 (0.8%) | 49 (6.6%) |
| MAOI | / | / | / | / | / | / | / | / |
| TCA | 103 (1.9%) | / | 99 (1.9%) | / | 101 (1.9%) | / | 128 (2.0%) | / |
| Atypical antidepressants | 550 (10.1%) | 58 (9.9%) | 494 (9.7%) | 46 (8.4%) | 538 (9.9%) | 55 (9.0%) | 654 (10.0%) | 62 (8.4%) |
| Task score | 2.6 ($\pm 0.8$) | 2.6 ($\pm 0.8$) | 0.4 ($\pm 0.2$) | 0.4 ($\pm 0.2$) | −5.2 ($\pm 1.8$) | −4.8 ($\pm 1.9$) | 0.9 ($\pm 0.1$) | 0.9 ($\pm 0.1$) |

Demographic-adjusted normative deviations *D* in MDD and BD groups were modelled using no-intercept regressions, such that each group’s regression coefficient *β* directly quantifies the standardized mean difference relative to healthy norms, equivalent to Δ_Glass_ = (*X̅*_group_ *X̅*_NCC_)*/s*_NCC_ [71]. Δ_Glass_ is conventionally interpreted against semi-qualitative thresholds of 0.2 (small), 0.5 (medium), and 0.8 (large).

Normative deviation (Figure 1 and Table 2) was significantly lower than zero for GradCPT in both MDD (*β* = 0.081, *q*_BH_ = 1.91 10^−8^) and BD (*β* = 0.187, *q*_BH_ = 6.70 10^−5^), indicating impaired sustained attention relative to NCC. Deviation was likewise significant for Delay Discounting in both MDD (*β* = +0.092, *q*_BH_ = 8.57 10^−10^) and BD (*β* = +0.240, *q*_BH_ = 3.08 10^−7^), showing an elevated preference for immediate over delayed rewards. The direct BD MDD contrast confirmed that BD participants exhibited greater impairment than MDD on both tasks. While the GradCPT contrast was significant only at the nominal level (*β* = 0.106, *p*_raw_ = 0.0168, *q*_BH_ = 0.0674), the Delay Discounting contrast survived joint FDR correction (*β* = +0.148, *q*_BH_ = 0.0063).

**Table 2:** Deviations (relative to NCC) in cognitive tasks across MDD and BD. Deviation *D* was modelled as *D* ∼ 0 + *C*(group). *β* is the raw coefficient from this model (with HC3 robust standard errors); since there is no intercept and *D* is expressed in units of NCC residual standard deviation, each group’s *β* coefficient equals that group’s mean *D* and is directly interpretable as an effect size (Δ_Glass_), representing how many standard deviations performance differs relative to demographically matched healthy norms. For the contrast row (BD − MDD), *β* is the Wald-test estimate of the difference between the two group coefficients from the same model. *p*_raw_ and *q*_BH_ are, respectively, the unadjusted two-sided *p*-value and its Benjamini-Hochberg-adjusted counterpart, computed jointly across all 28 tests in this study. grey-shaded rows denote *q*_BH_ *<* 0.05.

| Task | Group | MDD<br>(N) | BD<br>(N) | NCC<br>(N) | $\beta$ | 95% CI | $p_{\text{raw}}$ | $q_{\text{BH}}$ |
| --- | --- | --- | --- | --- | --- | --- | --- | --- |
| GradCPT | MDD | 5,472 | — | 43,515 | -0.081 | [-0.107, -0.055] | $1.362 \times 10^{-9}$ | $1.907 \times 10^{-8}$ |
| | BD | — | 584 | 43,515 | -0.187 | [-0.269, -0.104] | $9.569 \times 10^{-6}$ | $6.698 \times 10^{-5}$ |
|  | contrast | 5,472 | 584 | — | -0.106 | [-0.192, -0.019] | 0.01684 | 0.06735 |
| Flanker | MDD | 5,087 | — | 40,589 | -0.025 | [-0.053, +0.002] | 0.06769 | 0.2106 |
|  | BD | — | 545 | 40,589 | +0.033 | [-0.056, +0.123] | 0.4641 | 0.6213 |
|  | contrast | 5,087 | 545 | — | +0.059 | [-0.035, +0.152] | 0.2182 | 0.3819 |
| DD | MDD | 5,411 | — | 42,734 | +0.092 | [+0.065, +0.119] | $3.060 \times 10^{-11}$ | $8.569 \times 10^{-10}$ |
| | BD | — | 608 | 42,734 | +0.240 | [+0.155, +0.325] | $3.301 \times 10^{-08}$ | $3.081 \times 10^{-07}$ |
|  | contrast | 5,411 | 608 | — | +0.148 | [+0.059, +0.238] | 0.001128 | 0.006319 |
| EmoRecog | MDD | 6,536 | — | 51,491 | +0.035 | [+0.011, +0.058] | 0.003862 | 0.01802 |
|  | BD | — | 739 | 51,491 | +0.018 | [-0.053, +0.089] | 0.6259 | 0.7303 |
|  | contrast | 6,536 | 739 | — | -0.017 | [-0.092, +0.058] | 0.6556 | 0.7343 |

Our findings do not support a difference from the NCC normative population on Flanker in either MDD or BD, nor a significant BD MDD contrast for this task; none of these three tests was significant even at the nominal (uncorrected) level. For EmoRecog, MDD showed a significant deviation from NCC (*q_BH_* = 0.01802), though the magnitude of the MDD–NCC difference was tiny (+0.035). Neither the BD deviation (+0.018, *q_BH_* = 0.7303) nor the BD−MDD contrast (−0.017, *q_BH_* = 0.7343) reached significance for this task.

### 3.2 Association between wearable phenotypes and cognitive performance in MDD

The population of *EtM* completers had only partial overlap with the population of Fitbit users in the *All of Us* Research Program. Because this analysis investigates whether differences in wearable phenotypes associate with differences in cognitive deviation, the available sample was a subset of that used in the analysis above. The data retention cascade is reported in Supplementary Material (Table S3). Power analysis (Table S5c) revealed that the BD sample was powered to detect only correlations ranging from *r* = 0.297 (EmoRecog) to *r* = 0.323 (Flanker), unrealistically large for these phenotypes in a naturalistic epidemiological study; this limited power led us to exclude BD from this analysis.

Realised MDD sample sizes were 544 for GradCPT, 516 for Flanker, 538 for Delay Discounting, and 641 for EmoRecog. A sensitivity analysis (Table S6) showed that the MDD participants who provided wearable data did not differ systematically, on cognitive measures or covariates, from MDD participants who did not, suggesting the wearable-eligible subsample is representative of the broader MDD cohort rather than a distinct population. Clinical and demographic characteristics of the MDD sample used in this analysis are shown in Table 3.

**Table 3:** Analysis of associations between wearable phenotypes and cognitive task performance: Clinical-demographics of the MDD cohort across *EtM* tasks. Only a subset of *EtM* participants provided concurrent Fitbit data, resulting in a reduced sample size. As the restricted sample size yielded insufficient statistical power for the BD cohort, this analysis is restricted exclusively to the MDD group. The forward slash (/) indicates data suppressed to protect participant privacy in accordance with *All of Us* policies (*N <* 20). Prior episodes: Derived from EHR data using a heuristic that consolidated diagnostic entries within 180 days into a single episode. Abbreviations: CCI, Charlson Comorbidity Index (excluding mental health comorbidities); SSRI, Selective Serotonin Reuptake Inhibitor; SNRI, Serotonin-Norepinephrine Reuptake Inhibitor; Anticonvulsants (valproate, lamotrigine, carbamazepine); Atypical antidepressants (mirtazapine, trazodone, bupropion); MAOI, Monoamine Oxidase Inhibitor; TCA, Tricyclic Antidepressant. Medication counts reflect any prescriptions during the study follow-up period.

| Variable | GradCPT<br>($N = 544$) | Flanker<br>($N = 516$) | DD<br>($N = 538$) | EmoRecog<br>($N = 641$) |
| --- | --- | --- | --- | --- |
| Age (years) | 58.2 ($\pm 14.9$) | 58.2 ($\pm 15.0$) | 57.8 ($\pm 15.0$) | 57.5 ($\pm 14.7$) |
| Female sex | 409 (75.2%) | 387 (75.0%) | 402 (74.7%) | 479 (74.7%) |
| Education (0–7) | 5.9 ($\pm 1.0$) | 5.9 ($\pm 1.0$) | 5.9 ($\pm 1.0$) | 5.9 ($\pm 1.0$) |
| Touchscreen | 361 (66.4%) | 344 (66.7%) | 358 (66.5%) | 436 (68.0%) |
| BMI (kg/m<sup>2</sup>) | 31.1 ($\pm 7.9$) | 31.0 ($\pm 7.9$) | 30.9 ($\pm 7.9$) | 31.1 ($\pm 7.9$) |
| CCI | 3.2 ($\pm 3.8$) | 3.3 ($\pm 3.9$) | 3.2 ($\pm 3.8$) | 3.2 ($\pm 3.8$) |
| Season: |  |  |  |  |
| Spring | 6.4% | 5.8% | 7.1% | 7.0% |
| Summer | 16.5% | 17.4% | 14.5% | 16.5% |
| Autumn | 62.3% | 64.5% | 63.2% | 62.4% |
| Winter | 14.7% | 12.2% | 15.2% | 14.0% |
| Ethnicity: |  |  |  |  |
| White | 449 (82.5%) | 424 (82.2%) | 442 (82.2%) | 526 (82.1%) |
| Black | / | 20 (3.9%) | 23 (4.3%) | 30 (4.7%) |
| Asian | / | / | / | / |
| Medications: |  |  |  |  |
| SSRI | 71 (13.1%) | 67 (13.0%) | 71 (13.2%) | 83 (12.9%) |
| SNRI | 45 (8.3%) | 37 (7.2%) | 44 (8.2%) | 50 (7.8%) |
| Lithium | / | / | / | / |
| Antipsychotics | / | / | / | / |
| Anticonvulsants | / | / | / | / |
| MAOI | / | / | / | / |
| TCA | / | / | / | 22 (3.4%) |
| Atypical antidepressants | 53 (9.7%) | 47 (9.1%) | 45 (8.4%) | 64 (10.0%) |
| Prior episodes | 2.2 ($\pm 1.9$) | 2.2 ($\pm 1.9$) | 2.2 ($\pm 1.8$) | 2.2 ($\pm 1.8$) |
| Wearable Features: |  |  |  |  |
| Steps (mean/day) | 6,481.8 ($\pm 3,533.2$) | 6,600.6 ($\pm 3,347.6$) | 6,441.7 ($\pm 3,479.0$) | 6,538.1 ($\pm 3,378.3$) |
| WASO (min) | 55.0 ($\pm 12.9$) | 54.8 ($\pm 13.0$) | 54.8 ($\pm 13.1$) | 54.8 ($\pm 12.7$) |
| TST (min) | 389.2 ($\pm 58.2$) | 388.5 ($\pm 58.8$) | 388.7 ($\pm 60.5$) | 389.2 ($\pm 59.0$) |
| STV (h) | 1.3 ($\pm 0.6$) | 1.3 ($\pm 0.5$) | 1.3 ($\pm 0.5$) | 1.3 ($\pm 0.5$) |
| Task score | 2.6 ($\pm 0.8$) | 0.4 ($\pm 0.2$) | −5.3 ($\pm 1.8$) | 0.9 ($\pm 0.1$) |

None of the wearable phenotypes (Steps, WASO, TST, and STV) reached BH-corrected significance for any task (Table 4), and *R*^2^ was below 0.01 in every case. One test crossed the nominal (uncorrected) threshold (TST and EmoRecog at *p*_raw_ = 0.029) but did not survive correction (*q_BH_* = 0.103); no other test approached nominal significance. Given that this sample was powered to detect associations as small as *r* = 0.110 (*R*^2^ = 0.012) at nominal *p*-values (Table S5a), this near-total absence of even nominal signal indicates that wearable phenotypes sampled over the 90 days preceding testing show no meaningful association with cognitive performance in MDD, or that any true association is smaller than we had power to detect.

**Table 4:** Association between wearable phenotypes and cognitive deviation *D* in MDD. For each phenotype and task, each raw wearable phenotype was first residualised on age, sex, BMI, Charlson Comorbidity Index, and photoperiod, then standardised to zero mean and unit variance (*P_z_*; section 2.5.2). *D* was then regressed on *P_z_* (with HC3 robust standard errors): *D ∼ P_z_*. *β* is the estimated change in *D* per one unit change in *P_z_*. *R*^2^ is proportion of variance in *D* explained by *P_z_*; in simple linear (i.e., *D ∼ P_z_*) regression 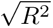 equals Pearson’s *r*. *p*_raw_ and *q*_BH_ are, respectively, the unadjusted two-sided *p*-value and its Benjamini-Hochberg-adjusted counterpart, computed jointly across all 28 tests in this study. grey-shaded rows denote *q*_BH_ *<* 0.05.

| Task | Fitbit Phenotype | $N$ | $\beta$ | 95% CI | $R^2$ | $p_{\text{raw}}$ | $q_{\text{BH}}$ |
| --- | --- | --- | --- | --- | --- | --- | --- |
| **GradCPT** | Steps | 544 | +0.056 | [-0.027, +0.139] | 0.0032 | 0.1868 | 0.3790 |
|  | WASO | 544 | +0.061 | [-0.027, +0.150] | 0.0038 | 0.1724 | 0.3790 |
|  | TST | 544 | +0.060 | [-0.032, +0.151] | 0.0036 | 0.2012 | 0.3790 |
|  | STV | 540 | -0.034 | [-0.133, +0.064] | 0.0012 | 0.4957 | 0.6213 |
| **Flanker** | Steps | 516 | +0.042 | [-0.038, +0.121] | 0.0016 | 0.3080 | 0.5008 |
|  | WASO | 516 | +0.079 | [-0.015, +0.172] | 0.0058 | 0.09874 | 0.2765 |
|  | TST | 516 | +0.033 | [-0.061, +0.126] | 0.0010 | 0.4918 | 0.6213 |
|  | STV | 512 | -0.037 | [-0.123, +0.050] | 0.0013 | 0.4037 | 0.5949 |
| **DD** | Steps | 538 | -0.075 | [-0.188, +0.037] | 0.0056 | 0.1879 | 0.3790 |
|  | WASO | 538 | +0.003 | [-0.079, +0.084] | 0.0000 | 0.9505 | 0.9505 |
|  | TST | 538 | -0.028 | [-0.112, +0.056] | 0.0008 | 0.5103 | 0.6213 |
|  | STV | 535 | +0.056 | [-0.030, +0.142] | 0.0031 | 0.2030 | 0.3790 |
| **EmoRecog** | Steps | 641 | -0.014 | [-0.084, +0.057] | 0.0002 | 0.7050 | 0.7481 |
|  | WASO | 641 | +0.013 | [-0.057, +0.083] | 0.0002 | 0.7213 | 0.7481 |
|  | TST | 641 | +0.076 | [+0.008, +0.145] | 0.0071 | 0.02946 | 0.1031 |
|  | STV | 636 | -0.036 | [-0.107, +0.035] | 0.0016 | 0.3220 | 0.5008 |

The sensitivity analysis in NCC indirectly supports this conclusion. For NCC, *D* was computed identically to MDD and BD: as the standardised residual from the same normative model, here applied to the population it was fitted on. Owing to its much larger sample size, the NCC sample was better powered to detect small effects than the MDD sample (Table S5b). Associations were in the expected direction: higher physical activity (Steps), longer TST, and lower STV were associated with better sustained attention (GradCPT) and lower impulsivity (DD), lower STV with better social cognition (EmoRecog). Flanker showed no association with any wearable phenotype.

A sensitivity analysis testing for a U-shaped relationship between TST and cognitive performance (Table S8, i.e., an optimal "sweet spot" with declining performance at both shorter and longer TSTs) found nominal, uncorrected evidence of such a pattern in the NCC sample for GradCPT (*p <* 0.001) and, marginally, EmoRecog (*p* = 0.025); no evidence was found in MDD for any task, or in NCC for Flanker or DD.

## 4 Discussion

This study employed data from the *All of Us* Research Program and asked two questions. First, we examined deviations in cognitive performance relative to NCC across MDD and BD in the three transdiagnostic cognitive domains of cold cognition (sustained attention, inhibitory control), hot cognition (reward-based impulsivity), and social cognition (facial emotion recognition). Second, in the MDD sample, we studied whether habitual, wearable-derived rest-activity phenotypes are associated with cognitive performance in across the aforementioned domains.

### 4.1 Cognitive performance across MDD, BD, and NCC

Both cold cognition, indexed by sustained attention on the GradCPT, and hot cognition, indexed by reward-based impulsivity on the Delay Discounting Task, differed significantly between each clinical group outside an acute mood episode and non-clinical controls. Both domains showed a consistent gradient of impairment, with euthymic BD associated with poorer sustained attention and steeper delay discounting than remitted MDD, which in turn performed worse than non-clinical controls (Table 2).

Persistent sustained-attention impairment has previously been reported in both euthymic BD and remitted MDD, although estimates vary considerably across tasks and performance indices [72, 73]. The present findings extend this literature by demonstrating, within the same large sample and using an identical assessment, an ordered gradient of poorer target discrimination from non-clinical controls to remitted MDD and euthymic BD.

Although meta-analytic evidence indicates steeper delay discounting in both MDD and BD, relatively few studies have instead examined inter-episode populations [74–76]. In remitted MDD, findings range from largely state-dependent alterations to small but significant persistent increases in discounting, whereas studies of clinically stable euthymic BD suggest steeper discounting and associations with poorer executive functioning [77–80].

Our findings support the hypothesis that both disorders involve alterations in hot cognition [14, 15]; however, steeper delay discounting in BD may distinguish it from MDD, potentially reflecting a stronger temperamental disposition towards reward-driven impulsivity [81]. Similarly, although impaired sustained attention appears to persist in both disorders, its greater severity in euthymic BD may distinguish it from remitted MDD, potentially reflecting a more pronounced trait-like vulnerability in attentional control [72, 73]. Interestingly, these two alterations may interact. A stronger bottom-up preference for immediate rewards, combined with weaker top-down attentional control, could make behaviour more strongly driven by salient short-term incentives and less effectively regulated by longer-term goals. In BD, this imbalance may amplify reward-driven activation and mood reactivity, thereby contributing to impulsive behaviour and potentially to a more recurrent or unstable illness course [34]. This interpretation is consistent with models linking hyperthymic temperament to immediate engagement with the environment and impulsive action, as well as with reward-hypersensitivity accounts of BD [81, 82]. Nevertheless, because the present tasks assessed these processes separately, their interaction remains a mechanistic hypothesis rather than a direct finding of the study.

In contrast to the group differences observed in sustained attention and delay discounting, attentional inhibition did not differ between non-clinical controls, remitted MDD and euthymic BD. This finding is consistent with evidence of preserved executive-attention networks in remitted MDD [83], although small studies have reported greater conflict interference in euthymic BD [84, 85]. This discrepancy may have two explanations. First, calculating the interference effect as a simple difference in accuracy may obscure alterations in the underlying decision-making process that could be detected using more fine-grained computational approaches [86]. Second, our sample comprised predominantly community-dwelling participants identified through electronic health records who completed the battery without supervision and may therefore have had milder illness or been better compensated than clinically recruited samples.

Although remitted MDD showed statistically higher accuracy than non-clinical controls, the magnitude of this difference was negligible, while the similarly small BD estimate did not reach significance and did not differ from that observed in MDD. Thus, these findings provide little evidence that facial-emotion recognition specifically distinguishes the two disorders; rather, the statistical significance of the MDD contrast likely reflects the substantially greater precision afforded by its larger sample. The absence of a detectable BD effect may also reflect differences between the present measure and the broader literature. Meta-analyses indicate that facial-emotion recognition deficits in euthymic BD are generally small [24–26], and may vary across specific emotions. By averaging accuracy across happiness, sadness, anger, and fear, our measure may have obscured emotion-specific alterations. Overall, the results do not support a broad facial-emotion labelling deficit during euthymia, but they cannot exclude subtler alterations in the processing of particular emotional expressions.

### 4.2 Rest-activity phenotypes and cognition in MDD

None of the four wearable phenotypes reached BH-corrected significance for any task in MDD, and no association explained more than 1% of variance (Table 4). This null is not attributable to inadequate power: MDES calculations show our sample was powered to detect associations as small as *r* 0.11–0.12 (Table S5), and the NCC sensitivity analysis, better powered still, detected several associations in the same *r* 0.05–0.11 range MDD would have struggled to resolve. So while our null does not rule out a small habitual rest-activity–cognition association of the magnitude seen in NCC, it does rule out effect sizes of clinical relevance.

This pattern is consistent with existing objective-measure evidence. A systematic review found most positive sleep– cognition associations in mood disorders came from subjective report, with objective (actigraphy, polysomnography) evidence sparse and inconsistent [40]; more recent objective-measure studies reporting positive associations have been conducted exclusively in BD and at far smaller scale (*N* = 34–40) [87, 88], leaving open whether they generalise to MDD or to habitual recording windows. A comparably large consumer-wearable MDD study reached the same null for physiologically measured sleep, despite self-reported sleep disturbance correlating with performance in the same sample [43], suggesting the signal may sit more in perceived sleep quality than in objective sleep parameters. It is also possible our phenotypes capture the wrong dimension of activity: cardiorespiratory fitness, rather than step volume, for example, has been shown to explain substantial variance in inhibitory control in MDD [89].

Sensitivity analyses lend further support to our findings as genuine null. A U-shaped TST–cognition relationship was detectable in the well-powered NCC sample but not in MDD for any task (Table S8), so a non-linearity missed by our primary linear analysis is unlikely to explain it. MDD participants included in versus excluded from the wearable analysis did not differ on cognitive deviation or clinical covariates (Table S6), arguing against selective loss of more impaired participants. BD was excluded from this analysis a priori for insufficient power (*N* = 73–87; Table S5c), so whether rest-activity phenotypes relate to cognition in BD, where we observed the largest cognitive deviations, remains open.

The near-total absence of association between wearable rest-activity phenotypes and cognitive performance in MDD is a sobering result. Given our sample size, the largest for studies of this kind to date and well powered to detect small effects, and the benchmark set by the NCC sensitivity analysis, this reads as a genuine null: consumer wearable metrics, step count, TST, WASO, and sleep timing variability, over a 90-day window, do not track cognitive functioning in MDD at an effect size of clinical relevance. Three caveats apply. The design is cross-sectional and observational, so we cannot say what would happen if rest-activity habits in MDD were deliberately changed. A single 90-day window may not represent an individual’s long-term habit history; if current patterns diverge from multi-year averages, this snapshot would miss the cumulative cognitive impact of chronic sleep or activity disruption. And our results speak only to the cognitive domains and rest-activity metrics measured here. Still, on this evidence, variation in the rest-activity phenotypes commonly offered by consumer devices does not track with performance in sustained attention, inhibitory control, reward-based impulsivity, or social cognition. This should temper expectations for how much a lifestyle intervention targeting sleep or activity alone would improve cognition in MDD.

### 4.3 Strengths

This study benefits from a significantly larger sample size than prior work examining either cognitive performance across euthymic MDD and BD relative to NCC, or the association between wearable rest-activity phenotypes and cognition in mood disorders, both of which have previously relied on samples in the tens to low hundreds [40, 87, 88]. This size gave us the power to detect effects considerably smaller than those reported in prior clinical literature, which we quantified directly by computing minimum detectable effect sizes for every planned test (Table S5). Prior work generally examined each disorder in isolation relative to healthy controls, whereas our study directly compares MDD and BD head-to-head within the same cohort. The RDoC-grounded, remotely administered EtM battery spans four dissociable transdiagnostic domains under one deployment protocol, and the NCC sensitivity analysis for the wearable-cognition association gave us an internal, well-powered benchmark against which to interpret the MDD null, rather than relying on external literature alone. The representativeness check and non-linearity check let us rule out two concrete alternative explanations for that null before treating it as informative.

### 4.4 Limitations

Several limitations should be taken into account. Participation in *All of Us*, completion of an unsupervised remote cognitive battery, and ownership or linkage of a Fitbit device jointly select for a population likely less impaired than clinically ascertained samples. Cognitive impairment, and any rest-activity–cognition association, may consequently be attenuated relative to what a clinical inpatient or specialist-referred sample would show.

The analysis is cross-sectional (most participants in data release CDRv9 - C2025Q4R6 completed the EtM battery just once), so we cannot distinguish stable trait-level cognitive deviation from slower within-person drift, of the kind a twelve-month actigraphy study was able to detect for depressive symptoms and attention [42]. Similarly, the 90-day Fitbit observation window strikes a pragmatic balance between capturing a representative rest-activity profile and maintaining a high sample size. However, this trade-off means the study cannot detect subtle or long-term signals that would require years of longitudinal tracking to observe.

Our four wearable phenotypes, step count, TST, WASO, and sleep timing variability, do not capture sleep architecture, autonomic measures, or activity intensity and fitness, the last of which has itself been linked to inhibitory control in MDD independently of step volume [89]. We also lacked a subjective sleep-quality measure, so we cannot directly test the objective-subjective discordance reported elsewhere [43]. While shift-work status (a confounder of rest-activity phenotypes) was not explicitly recorded in our dataset, our quality control procedures (excluding sleep episodes initiating outside 18:00–08:00) partially mitigated this issue.

On the cognitive tests’ side, Flanker and EmoRecog each yield a single composite score rather than a fuller characterisation. For Flanker, a metric more sensitive to intra-individual response-time variability might behave differently, as discussed above. For EmoRecog, averaging accuracy across all four emotions cannot speak to whether recognition of specific, particularly negative, emotions is selectively affected in MDD or BD [90]; a genuine valence-specific impairment could in principle coexist with the small overall positive deviation (MDD) or null (BD) reported here. The EtM battery does not assess memory or processing speed, two domains that meta-analytic evidence identifies among the more consistently, and in some subgroups the most severely, affected in euthymic MDD [19].

BD was excluded from the wearable-cognition analysis due to statistical power considerations (Section 2.6; Table S5c), leaving this question unaddressed in the group that showed the larger cognitive deviations in this study.

Medication exposure differed substantially between MDD and BD, for example antipsychotic use at 1.4% versus 7.7% and anticonvulsant use at 0.7% versus 6.7% (Table 1), and was not adjusted for in the group contrasts; some portion of the BD*>*MDD cognitive gradient may reflect medication exposure rather than illness-specific severity. Disease onset was left-censored for many participants, and too few had recorded psychotic episodes to examine either as a correlate of cognitive performance, likely reflecting further under-representation of the most severely affected end of the clinical spectrum in this epidemiological sample.

### 4.5 Conclusions

In conclusion, remitted MDD and euthymic BD were characterised by selective rather than generalised cognitive alterations. Sustained attention and delay discounting showed a consistent gradient from non-clinical controls to remitted MDD and euthymic BD, whereas attentional inhibition and overall facial-emotion recognition showed no clinically meaningful group differences. These findings indicate that symptomatic remission does not necessarily entail cognitive recovery and support targeted assessment of sustained attentional control and reward-based decision-making, particularly in BD. Conversely, the null findings caution against assuming pervasive impairment across all cognitive domains or interpreting statistically detectable but negligible differences as clinically relevant.

Wearable-derived step count, sleep duration, wake after sleep onset, and sleep-timing variability did not meaningfully explain cognitive heterogeneity within remitted MDD. Accordingly, these commonly available consumer-wearable measures should not presently be used as proxies for cognitive functioning or to identify individuals requiring cognitive intervention. This does not diminish the broader physical and mental health benefits of improving sleep and activity, nor does the cross-sectional design exclude cognitive benefits from experimentally modifying these behaviours. Rather, the findings suggest that persistent cognitive difficulties require direct, domain-specific assessment and that wearable markers with greater mechanistic proximity may be needed to capture clinically informative relationships.

## Data Availability

Data were sourced from the All of Us Research Program and are not owned by the authors. Access protocols and eligibility criteria can be found at: https://support.researchallofus.org/hc/en-us/articles/22346942074132-Data-Access-Framework

## 5 Acknowledgments

We gratefully acknowledge *All of Us* participants for their contributions, without whom this research would not have been possible. We also thank the National Institutes of Health’s All of Us Research Program for making available the participant data examined in this study.

## 6 Authors contributions

**F.C.:** Conceptualisation, Methodology, Code Implementation, Writing; **M.K.:** Conceptualisation, Writing; **M.R.:** Writing **P.O.:** Conceptualisation, Writing; **G.F.:** Reviewing; **S.J.:** Writing; **A.H.Y.:** Reviewing. All authors contributed substantially to the work, read, and approved the final manuscript.

## 8 Data and Code Availability

Data were sourced from the *All of Us* Research Program and are not owned by the authors. Access protocols and eligibility criteria can be found at the All of Us Research Program Support Portal. The complete code repository required to reproduce the analysis presented in this paper will be made publicly available upon peer-reviewed publication at https://github.com/FilippoCMC.

## 9 Ethical considerations

Authorised researchers performed a secondary analysis of de-identified *All of Us* Research Program data within the Verily Researcher Workbench. Institutional Review Board approval was not required because the study relied exclusively on existing de-identified data with no participant contact. Informed consent was obtained from all participants at enrollment.

## 10 AI use disclosure

The authors used Anthropic’s Claude Sonnet 5 for language editing and improvement of sentence formulation. No scientific content, interpretation, literature selection, or conclusions were generated by the tool. The authors critically reviewed and revised all AI-assisted text and take full responsibility for the accuracy, interpretation, and final wording of the manuscript.

## References

[1] Andre F Carvalho, Joseph Firth, and Eduard Vieta. Bipolar disorder. New England Journal of Medicine, 383(1): 58–66, 2020.

[2] Wolfgang Marx, Brenda WJH Penninx, Marco Solmi, Toshi A Furukawa, Joseph Firth, Andre F Carvalho, and Michael Berk. Major depressive disorder. Nature Reviews Disease Primers, 9(1):44, 2023.

[3] F. Hardeveld, J. Spijker, R. De Graaf, W. A. Nolen, and A. T. F. Beekman. Prevalence and predictors of recurrence of major depressive disorder in the adult population. Acta Psychiatrica Scandinavica, 122(3):184–191, 2010. doi: 10.1111/j.1600-0447.2009.01519.x.

[4] Andreanne Gignac, Alexander McGirr, Raymond W Lam, and Lakshmi N Yatham. Recovery and recurrence following a first episode of mania: a systematic review and meta-analysis of prospectively characterized cohorts. The Journal of clinical psychiatry, 76(9):1241–1248, 2015.

[5] Lene Hammer-Helmich, Josep Maria Haro, Bengt Jönsson, Audrey Tanguy Melac, Sylvie Di Nicola, Julien Chollet, Dominique Milea, Benoît Rive, and Delphine Saragoussi. Functional impairment in patients with major depressive disorder: The 2-year perform study. Neuropsychiatric Disease and Treatment, 14:239–249, 2018. doi: 10.2147/NDT.S146098.

[6] Ronald C. Kessler, Patricia Berglund, Olga Demler, Robert Jin, Doreen Koretz, Kathleen R. Merikangas, A. John Rush, Ellen E. Walters, and Philip S. Wang. The epidemiology of major depressive disorder: Results from the national comorbidity survey replication (ncs-r). JAMA, 289(23):3095–3105, 2003. doi: 10.1001/jama.289.23.3095.

[7] Gabriela Léda-Rêgo, Severino Bezerra-Filho, and Ângela Miranda-Scippa. Functioning in euthymic patients with bipolar disorder: A systematic review and meta-analysis using the functioning assessment short test. Bipolar Disorders, 22(6):569–581, 2020. doi: 10.1111/bdi.12904.

[8] Dimosthenis Tsapekos, Michail Kalfas, Rebecca Strawbridge, Samuel Swidzinski, Katherine E. Burdick, and Allan H. Young. Estimating cognitive impairment in bipolar disorder: Should we account for premorbid iq? Acta Psychiatrica Scandinavica, 153(5):468–476, 2025. doi: 10.1111/acps.70000.

[9] Tanya Tran, Melissa Milanovic, Katherine Holshausen, and Christopher R. Bowie. What is normal cognition in depression? prevalence and functional correlates of normative versus idiographic cognitive impairment. Neuropsychology, 35(1):33–41, 2021. doi: 10.1037/neu0000717.

[10] Maria Semkovska, Lisa Quinlivan, Tara O’Grady, Rebecca Johnson, Aisling Collins, Jessica O’Connor, Hannah Knittle, Elayne Ahern, and Taylor Gload. Cognitive function following a major depressive episode: A systematic review and meta-analysis. The Lancet Psychiatry, 6(10):851–861, 2019. doi: 10.1016/S2215-0366(19)30291-3.

[11] Johan Høy Jensen, Ulla Knorr, Maj Vinberg, Lars V. Kessing, and Kamilla W. Miskowiak. Discrete neurocognitive subgroups in fully or partially remitted bipolar disorder: Associations with functional abilities. Journal of Affective Disorders, 205:378–386, 2016. doi: 10.1016/j.jad.2016.08.018.

[12] Mickael Ehrminger, Eric Brunet-Gouet, Anne-Sophie Cannavo, Bruno Aouizerate, Irena Cussac, Jean-Michel Azorin, Frank Bellivier, Thierry Bougerol, Philippe Courtet, Caroline Dubertret, Bruno Etain, Jean-Pierre Kahn, Marion Leboyer, Emilie Olié, Christine Passerieux, and Paul Roux. Longitudinal relationships between cognition and functioning over 2 years in euthymic patients with bipolar disorder: A cross-lagged panel model approach with the face-bd cohort. The British Journal of Psychiatry, 218(2):80–87, 2021. doi: 10.1192/bjp.2019.180.

[13] Vanessa C. Evans, Grant L. Iverson, Lakshmi N. Yatham, and Raymond W. Lam. The relationship between neurocognitive and psychosocial functioning in major depressive disorder: A systematic review. The Journal of Clinical Psychiatry, 75(12):1359–1370, 2014. doi: 10.4088/JCP.13r08939.

[14] Jonathan P Roiser and Barbara J Sahakian. Hot and cold cognition in depression. CNS spectrums, 18(3):139–149, 2013.

[15] Tamsym Elizabeth Van Rheenen, Kathryn E Lewandowski, Amy Pinkham, Cristina Varo, Georgia Caruana, June Gruber, Jeff Zarp, Allan H Young, Lakshmi N Yatham, E Vieta, et al. Consensus on subdomains and measures of relevance to affective and social cognition research on bipolar disorder (cas-bd); outcomes and recommendations from an international society for bipolar disorders targeting cognition taskforce study. Bipolar disorders, 28(2): e70083, 2026.

[16] Margaret A Niznikiewicz. The building blocks of social communication. Advances in Cognitive Psychology, 9(4): 173, 2013.

[17] C Bourne, ÖMER Aydemir, Vicent Balanzá-Martínez, Emre Bora, Sofia Brissos, JTO Cavanagh, Luke Clark, Zeynep Cubukcuoglu, Vasco Videira Dias, Sandra Dittmann, et al. Neuropsychological testing of cognitive impairment in euthymic bipolar disorder: an individual patient data meta-analysis. Acta Psychiatrica Scandinavica, 128(3):149–162, 2013.

[18] Samuel Swidzinski, Dimosthenis Tsapekos, Pricilla Swidzinska, Wenjia Zhang, Moxun Zheng, Edward Millgate, Rebecca Strawbridge, Roxanna Short, Ben Carter, Peter Gallagher, et al. Domain-specific cognitive function in euthymic bipolar disorder: a systematic review and meta-analysis. Psychological Medicine, 55:e336, 2025.

[19] E. Bora, B. J. Harrison, M. Yücel, and C. Pantelis. Cognitive impairment in euthymic major depressive disorder: a meta-analysis. Psychological Medicine, 43(10):2017–2026, oct 2013. doi: 10.1017/S0033291712002085.

[20] Dominik Kriesche, Christian FJ Woll, Nadja Tschentscher, Rolf R Engel, and Susanne Karch. Neurocognitive deficits in depression: a systematic review of cognitive impairment in the acute and remitted state. European archives of psychiatry and clinical neuroscience, 273(5):1105–1128, 2023.

[21] Elayne Ahern, Jessica White, and Eadaoin Slattery. Change in cognitive function over the course of major depressive disorder: A systematic review and meta-analysis. Neuropsychology Review, 35(1):1–34, 2025.

[22] Kamilla W Miskowiak, Ida Seeberg, Hanne L Kjaerstad, Katherine E Burdick, Anabel Martinez-Aran, Caterina del Mar Bonnin, Christopher R Bowie, Andre F Carvalho, Peter Gallagher, Gregor Hasler, et al. Affective cognition in bipolar disorder: a systematic review by the isbd targeting cognition task force. Bipolar disorders, 21 (8):686–719, 2019.

[23] Kamilla W. Miskowiak and Andre F. Carvalho. ‘hot’cognition in major depressive disorder: A systematic review. CNS & Neurological Disorders-Drug Targets-CNS & Neurological Disorders), 13(10):1787–1803, 2014.

[24] Cecilia Samame, Diego Javier Martino, and SA Strejilevich. Social cognition in euthymic bipolar disorder: Systematic review and meta-analytic approach. Acta Psychiatrica Scandinavica, 125(4):266–280, 2012.

[25] Emily S Gillissie, Leanna MW Lui, Felicia Ceban, Kamilla Miskowiak, Sena Gok, Bing Cao, Kayla M Teopiz, Roger Ho, Yena Lee, Joshua D Rosenblat, et al. Deficits of social cognition in bipolar disorder: systematic review and meta-analysis. Bipolar Disorders, 24(2):137–148, 2022.

[26] Michele De Prisco, Vincenzo Oliva, Chiara Possidente, Giovanna Fico, Laura Montejo, Lydia Fortea, Hanne Lie Kjærstad, Kamilla Woznica Miskowiak, Gerard Anmella, Diego Hidalgo-Mazzei, Alessandro Miola, Michele Fornaro, Andrea Murru, Eduard Vieta, and Joaquim Radua. Facial emotion recognition deficits in bipolar disorder: A systematic review and meta-analysis. European Psychiatry, 69(1):e16, jan 2026. doi: 10.1192/j.eurpsy.2025.10147.

[27] Michael James Weightman, Tracy Michele Air, and Bernhard Theodor Baune. A review of the role of social cognition in major depressive disorder. Frontiers in psychiatry, 5:179, 2014.

[28] Alainna Wen, Ethan Ray Fischer, David Watson, and K Lira Yoon. Biased cognitive control of emotional information in remitted depression: A meta-analytic review. Journal of psychopathology and clinical science, 132 (8):921, 2023.

[29] Cecilia Samame, AG Szmulewicz, Marina Paula Valerio, Diego Javier Martino, and SA Strejilevich. Are major depression and bipolar disorder neuropsychologically distinct? a meta-analysis of comparative studies. European Psychiatry, 39:17–26, 2017.

[30] Alejandro G Szmulewicz, Marina P Valerio, José M Smith, Cecilia Samamé, Diego J Martino, and Sergio A Strejilevich. Neuropsychological profiles of major depressive disorder and bipolar disorder during euthymia. a systematic literature review of comparative studies. Psychiatry research, 248:127–133, 2017.

[31] Kate Eggleston, Kamilla Woznica Miskowiak, Richard Porter, Chris Frampton, and Katie Douglas. Systematic review and meta-analysis of the association between subjective and objective cognitive function in mood disorders. Bipolar disorders, 28(1):e70077, 2026.

[32] Quentin JM Huys and Michael Browning. A computational view on the nature of reward and value in anhedonia. Current topics in behavioral neurosciences, 58:421–442, 2022.

[33] Catherine J Harmer and Michael Browning. Can a predictive processing framework improve the specification of negative bias in depression? Biological Psychiatry, 87(5):382–383, 2020.

[34] Liam Mason, Eran Eldar, and Robb B Rutledge. Mood instability and reward dysregulation—a neurocomputational model of bipolar disorder. JAMA psychiatry, 74(12):1275–1276, 2017.

[35] Michael James Weightman, Matthew James Knight, and Bernhard Theodor Baune. A systematic review of the impact of social cognitive deficits on psychosocial functioning in major depressive disorder and opportunities for therapeutic intervention. Psychiatry research, 274:195–212, 2019.

[36] Kamilla W Miskowiak, Ida Seeberg, Mette B Jensen, Vicent Balanza-Martinez, Caterina del Mar Bonnin, Christopher R Bowie, Andre F Carvalho, Annemieke Dols, Katie Douglas, Peter Gallagher, et al. Randomised controlled cognition trials in remitted patients with mood disorders published between 2015 and 2021: A systematic review by the international society for bipolar disorders targeting cognition task force. Bipolar disorders, 24(4): 354–374, 2022.

[37] Gin S Malhi, Lauren Irwin, Amber Hamilton, Grace Morris, Philip Boyce, Roger Mulder, and Richard J Porter. Modelling mood disorders: an ace solution? Bipolar disorders, 20:4–16, 2018.

[38] Ian B Hickie, Kathleen R Merikangas, Joanne S Carpenter, Frank Iorfino, Elizabeth M Scott, Jan Scott, and Jacob J Crouse. Does circadian dysrhythmia drive the switch into high-or low-activation states in bipolar i disorder? Bipolar Disorders, 25(3):191–199, 2023.

[39] Kathleen Ries Merikangas, Joel Swendsen, Ian B Hickie, Lihong Cui, Haochang Shou, Alison K Merikangas, Jihui Zhang, Femke Lamers, Ciprian Crainiceanu, Nora D Volkow, et al. Real-time mobile monitoring of the dynamic associations among motor activity, energy, mood, and sleep in adults with bipolar disorder. JAMA psychiatry, 76(2):190–198, 2019.

[40] Oliver Pearson, Nora Uglik-Marucha, Kamilla W. Miskowiak, Scott A. Cairney, Ivana Rosenzweig, Allan H. Young, and Paul R. A. Stokes. The relationship between sleep disturbance and cognitive impairment in mood disorders: A systematic review. Journal of Affective Disorders, 327:207–216, apr 2023. doi: 10.1016/j.jad.2023. 01.114.

[41] Kristine A Wilckens, Christopher E Kline, Marissa A Bowman, Ryan C Brindle, Matthew R Cribbet, Julian F Thayer, and Martica H Hall. Does objectively-assessed sleep moderate the association between history of major depressive disorder and task-switching? Journal of affective disorders, 265:216–223, 2020.

[42] Hang-Ju Yang, Wan-Ju Cheng, Mi-Chun Hsiao, Sheng-Che Huang, Tomohide Kubo, Liang-Wen Hang, and Wei-Sheng Lee. Rest–activity rhythm associated with depressive symptom severity and attention among patients with major depressive disorder: a 12-month follow-up study. Frontiers in Psychiatry, 14:1214143, aug 2023. doi: 10.3389/fpsyt.2023.1214143.

[43] Samir Akre, Zachary D Cohen, Amelia Welborn, Tomislav D Zbozinek, Brunilda Balliu, Michelle G Craske, and Alex AT Bui. Comparing self reported and physiological sleep quality from consumer devices to depression and neurocognitive performance. NPJ digital medicine, 8(1):92, 2025.

[44] Florian Wüthrich, Carver B Nabb, Vijay A Mittal, Stewart A Shankman, and Sebastian Walther. Actigraphically measured psychomotor slowing in depression: systematic review and meta-analysis. Psychological medicine, 52 (7):1208–1221, 2022.

[45] Fiona Yan-Yee Ho, Chun-Yin Poon, Vincent Wing-Hei Wong, Ka-Wai Chan, Ka-Wai Law, Wing-Fai Yeung, and Ka-Fai Chung. Actigraphic monitoring of sleep and circadian rest-activity rhythm in individuals with major depressive disorder or depressive symptoms: a meta-analysis. Journal of Affective Disorders, 361:224–244, 2024.

[46] Priyanka Panchal, Gabriela de Queiroz Campos, Danielle A Goldman, Randy P Auerbach, Kathleen R Merikangas, Holly A Swartz, Anjali Sankar, and Hilary P Blumberg. Toward a digital future in bipolar disorder assessment: a systematic review of disruptions in the rest-activity cycle as measured by actigraphy. Frontiers in psychiatry, 13: 780726, 2022.

[47] All of Us Research Program Investigators. The “all of us” research program. New England Journal of Medicine, 381(7):668–676, 2019.

[48] All of Us Research Program. Overview of exploring the mind data in the researcher workbench. All of Us Research Hub Support Documentation, 2026. URL https://support.researchallofus.org/hc/en-us/articles/33451505815316-Overview-of-Exploring-the-Mind-Data-in-the-Researcher-Workbench. Accessed: July 2026.

[49] Bruce N Cuthbert and Thomas R Insel. Toward the future of psychiatric diagnosis: the seven pillars of rdoc. BMC medicine, 11(1):1–8, 2013.

[50] Bruce N Cuthbert. Research domain criteria (rdoc): Progress and potential. Current directions in psychological science, 31(2):107–114, 2022.

[51] Laura Germine, Ken Nakayama, Bradley C Duchaine, Christopher F Chabris, Garga Chatterjee, and Jeremy B Wilmer. Is the web as good as the lab? comparable performance from web and lab in cognitive/perceptual experiments. Psychonomic bulletin & review, 19(5):847–857, 2012.

[52] Shifali Singh, Roger W Strong, Laneé Jung, Frances Haofei Li, Liz Grinspoon, Luke S Scheuer, Eliza J Passell, Paolo Martini, Naomi Chaytor, Jason R Soble, et al. The testmybrain digital neuropsychology toolkit: Development and psychometric characteristics. Journal of Clinical and Experimental Neuropsychology, 43(8):786–795, 2021.

[53] Jeffrey G Klann, Matthew AH Joss, Kevin Embree, and Shawn N Murphy. Data model harmonization for the all of us research program: Transforming i2b2 data into the omop common data model. PloS one, 14(2):e0212463, 2019.

[54] Joshua J Matacotta, Derek Tran, and Sonyeol Yoon. The prevalence of major depressive disorder in people with hiv: Results from the all of us research program. HIV medicine, 25(8):998–1004, 2024.

[55] Bhaavyaa B Shah, Michael L Thomas, Michael J McCarthy, and Alejandro D Meruelo. Body mass index mediates the relationship between depression and triglyceride levels: Evidence from a large national cohort. Journal of affective disorders, 390:119889, 2025.

[56] Yue Hu, Menglu Che, and Heping Zhang. Sex-specific association between polymorphisms in estrogen receptor alpha gene (esr1) and depression: A genome-wide association study of all of us and uk biobank data. Genetic epidemiology, 49(3):e70004, 2025.

[57] All of Us Research Program. All of Us Controlled Tier Dataset v9 CDR Data Dictionary (C2025Q4R6). Google Sheets Spreadsheet, 2025. URL https://docs.google.com/spreadsheets/d/1GKvrdnvOfzFW4yFveJL__c5mlZk_y6o7Pn8-8DZyYWs/edit?gid=1460903436#gid=1460903436. Table/Sheet: Exploring the Mind.

[58] Hiral Master, Jeffrey Annis, Shi Huang, Joshua A Beckman, Francis Ratsimbazafy, Kayla Marginean, Robert Carroll, Karthik Natarajan, Frank E Harrell, Dan M Roden, et al. Association of step counts over time with the risk of chronic disease in the all of us research program. Nature medicine, 28(11):2301–2308, 2022.

[59] Salim Yakdan, Braeden Benedict, Pranay Singh, Madelyn R Frumkin, Burel R Goodin, Brian Neuman, Abby L Cheng, Jing Wang, Michael P Kelly, Wilson Z Ray, et al. Association of activity with the risk of developing musculoskeletal pain in the all of us research program. The journal of pain, page 105516, 2025.

[60] Neil S Zheng, Jeffrey Annis, Hiral Master, Lide Han, Karla Gleichauf, Jack H Ching, Melody Nasser, Peyton Coleman, Stacy Desine, Douglas M Ruderfer, et al. Sleep patterns and risk of chronic disease as measured by long-term monitoring with commercial wearable devices in the all of us research program. Nature medicine, 30 (9):2648–2656, 2024.

[61] All of Us Research Program. Resources for using Fitbit data, 2025. URL https://support.researchallofus.org/hc/en-us/articles/20281023493908-Resources-for-Using-Fitbit-Data. Accessed: March 23, 2026.

[62] Andre C Tonon, Adile Nexha, Jasmyn EA Cunningham, Jason d’Eon, Trisha Chakrabarty, Faranak Farzan, Jane A Foster, Kate L Harkness, Stefanie Hassel, Keith Ho, et al. One-year actigraphy study of sleep and rest-activity rhythms as markers of relapse in depression. JAMA psychiatry, 83(4):379–388, 2026.

[63] Eliza Passell, Roger W Strong, Lauren A Rutter, Heesu Kim, Luke Scheuer, Paolo Martini, Liz Grinspoon, and Laura Germine. Cognitive test scores vary with choice of personal digital device. Behavior Research Methods, 53 (6):2544–2557, 2021.

[64] Hude Quan, Bing Li, Chantal M Couris, Kiyohide Fushimi, Patrick Graham, Phil Hider, Jean-Marie Januel, and Vijaya Sundararajan. Updating and validating the charlson comorbidity index and score for risk adjustment in hospital discharge abstracts using data from 6 countries. American journal of epidemiology, 173(6):676–682, 2011.

[65] J. W. Spencer. Fourier series representation of the position of the sun. Search, 2(5):162–172, 1971.

[66] J Almorox, C Hontoria, and M Benito. Statistical validation of daylength definitions for estimation of global solar radiation in toledo, spain. Energy Conversion and Management, 46(9-10):1465–1471, 2005.

[67] Eike Luedeling, Lars Caspersen, and Eduardo Fernandez. chillR: Statistical Methods for Phenology Analysis in Temperate Fruit Trees, 2025. URL https://rdrr.io/cran/chillR/src/R/daylength.R. R package version 0.77.

[68] Roman Yurchak and contributors. pgeocode: Postal code geocoding and distance calculation in python, 2024. URL https://pypi.org/project/pgeocode/. Maintained by Symerio. Available at https://github.com/symerio/pgeocode.

[69] John M Hoenig and Dennis M Heisey. The abuse of power: the pervasive fallacy of power calculations for data analysis. The American Statistician, 55(1):19–24, 2001.

[70] Alen Juginović. Balancing between too little and too much sleep. In Sleep Science Made Simple: A Clear and Concise Guide, pages 99–100. Springer Nature Switzerland, Cham, 2025. ISBN 978-3-031-89304-9.

[71] Gene V Glass, Barry McGaw, and Mary Lee Smith. Meta-analysis in social research. (No Title), 1981.

[72] Luke Clark and Guy M Goodwin. State-and trait-related deficits in sustained attention in bipolar disorder. European archives of psychiatry and clinical neuroscience, 254(2):61–68, 2004.

[73] Bethany Little, Megan Anwyll, Laura Norsworthy, Luke Corbett, Mia Schultz-Froggatt, and Peter Gallagher. Processing speed and sustained attention in bipolar disorder and major depressive disorder: A systematic review and meta-analysis. Bipolar Disorders, 26(2):109–128, mar 2024. doi: 10.1111/bdi.13396.

[74] Michael Amlung, Emma Marsden, Katherine Holshausen, Vanessa Morris, Herry Patel, Lana Vedelago, Katherine R Naish, Derek D Reed, and Randi E McCabe. Delay discounting as a transdiagnostic process in psychiatric disorders: A meta-analysis. JAMA Psychiatry, 76(11):1176–1186, nov 2019. doi: 10.1001/jamapsychiatry.2019.2102.

[75] Luis Felipe Sarmiento, Jorge Alexander Ríos-Flórez, Hector Andres Paez-Ardila, Pêssi Socorro Lima de Sousa, Antonio Olivera-La Rosa, Anderson Manoel Herculano Oliveira da Silva, and Amauri Gouveia Jr. Pharmacological modulation of temporal discounting: a systematic review. In Healthcare, volume 11, page 1046. MDPI, 2023.

[76] Qianying Li, Xiaofang Zhang, and Zhiguo Hu. Delay discounting in emotional disorders: A three-level meta-analysis. Journal of Affective Disorders, page 120515, 2025.

[77] E Pulcu, PD Trotter, EJ Thomas, M McFarquhar, Gabriella Juhász, BJ Sahakian, JFW Deakin, R Zahn, IM Anderson, and R Elliott. Temporal discounting in major depressive disorder. Psychological medicine, 44(9):1825–1834, 2014.

[78] Woo-Young Ahn, Olga Rass, Daniel J Fridberg, Anthony J Bishara, Jennifer K Forsyth, Alan Breier, Jerome R Busemeyer, William P Hetrick, Amanda R Bolbecker, and Brian F O’Donnell. Temporal discounting of rewards in patients with bipolar disorder and schizophrenia. Journal of abnormal psychology, 120(4):911, 2011.

[79] Alexandra K Gold and Michael W Otto. Why now and not later? an exploration into the neurocognitive correlates of delay discounting in bipolar disorder. Psychiatry research communications, 3(2):100114, 2023.

[80] Doron Elad, Giles W Story, Isabel M Berwian, Klaas E Stephan, Henrik Walter, and Quentin JM Huys. Delay discounting correlates with depression but does not predict relapse after antidepressant discontinuation. Molecular Psychiatry, 31(5):2445–2453, 2026.

[81] Hazel Tingzhu Chen, Matteo Martino, Elham Dabiri, Frans Ricardo Tamara, Lungile Sibiya, Benedetta Conio, Mario Amore, Thierry Burnouf, and Paola Magioncalda. Biological correlates of temperament: systematic reviews, empirical studies, and a conceptual framework linking neurotransmitter signaling, intrinsic brain activity, and the hyperthymic-depressive spectrum. Molecular Psychiatry, 30(12):5880–5888, 2025.

[82] Lauren B Alloy, Thomas Olino, Rachel D Freed, and Robin Nusslock. Role of reward sensitivity and processing in major depressive and bipolar spectrum disorders. Behavior therapy, 47(5):600–621, 2016.

[83] Kai-Jie Liang, Chun-Che Hung, Chia-Wen Ko, Pin-Chi Lin, Pei-Ying S Chan, and Chia-Hsiung Cheng. Error processing in major depressive disorder: a systematic review and meta-analysis of event-related potential studies. Clinical Neurophysiology, page 2111503, 2026.

[84] M. Preiss, L. Kramska, E. Dockalova, M. Holubova, and H. Kucerova. Attentional networks in euthymic patients with unipolar depression. European Psychiatry, 25(2):69–74, mar 2010. doi: 10.1016/j.eurpsy.2009.08.002.

[85] Andrea Marotta, Roberto Delle Chiaie, Alfredo Spagna, Laura Bernabei, Martina Sciarretta, Javier Roca, Massimo Biondi, and Maria Casagrande. Impaired conflict resolution and vigilance in euthymic bipolar disorder. Psychiatry research, 229(1-2):490–496, 2015.

[86] Paolo Ossola, Camilla Antonucci, Kevin B Meehan, Nicole M Cain, Martina Ferrari, Antonio Soliani, Carlo Marchesi, John F Clarkin, Fabio Sambataro, and Chiara De Panfilis. Effortful control is associated with executive attention: A computational study. Journal of personality, 89(4):774–785, 2021.

[87] Anna Tröger, Jules Schneider, Dimosthenis Tsapekos, Stephan Bialonski, Niklas Grieger, Michail Kalfas, Lisa Frending, Hannah Hartland, Heidi Kuivaniemi-Smith, Allan H. Young, Philipp Ritter, and Rebecca Strawbridge. The association between sleep spindles and cognitive performance in euthymic bipolar disorder. Translational Psychiatry, 16(1), jul 2026. doi: 10.1038/s41398-026-04273-2.

[88] Candice Libourel, Mathilde Charron, Victoire Martinot, Mathilde Carminati, Bruno Etain, and Vincent Hennion. Objective sleep parameters and cognitive performance in euthymic bipolar disorder: a cross-sectional 21-day actigraphy study. International Journal of Bipolar Disorders, 2026.

[89] Nils H. Pixa, Stephanie Fröhlich, Tim Göcking, Lothar Thorwesten, Sarah E. Fromme, Bernhard T. Baune, and Claudia Voelcker-Rehage. A cross-sectional study on the relationship between cardiorespiratory fitness, inhibitory control, and event-related potentials moderated by severity of symptoms in patients with major depressive disorder. Journal of Affective Disorders, 389:119701, nov 2025. doi: 10.1016/j.jad.2025.119701.

[90] Marta Monferrer, Arturo S. García, Jorge J. Ricarte, María J. Montes, Antonio Fernández-Caballero, and Patricia Fernández-Sotos. Facial emotion recognition in patients with depression compared to healthy controls when using human avatars. Scientific Reports, 13:6007, apr 2023. doi: 10.1038/s41598-023-31277-5.

