## Supplementary Material for "Transdiagnostic Domain-specific Cognitive Impairment in Inter-episode Mood Disorders and its Relationship with Rest-Activity Phenotypes"

Table S1: **Quality control flags applied to *Exploring the Mind* task sessions**. A task session was excluded if any exclusion criteria were true (see *All of Us* Research Program).

| Task | Primary score | Direction | Exclusion criterion |
|---|---|---|---|
| GradCPT | $d'$ | Higher = better attention | $\geq$75 consecutive non-responses ($\sim$60 s) |
| | | | $> 50\%$ omissions on go trials |
| | | | Cumulative flagged trial duration $> 60$ s |
| Flanker | $\mathrm{RCS}_{\mathrm{int}}$ | Higher = worse control | Overall accuracy $< 0.65$ |
| | | | $> 10\%$ of trials with RT $< 200$ ms |
| DD | $\ln k$ | Higher = more impulsive | Catch trial accuracy $< 0.75$ |
| | | | Median RT $< 500$ ms |
| EmoRecog | Avg. Accuracy | Higher = better | Median correct RT $< 700$ ms |
| | | | Any single response used on $> 90\%$ of trials |
| | | | $> 10\%$ of trials with RT $< 300$ ms |

Table S2: **OMOP concept IDs used for psychiatric diagnosis ascertainment and cohort (mutually exclusive, MDD, BD, or NCC) assignment**.

| Concept ID | Concept Name | Role in Cohort Assembly |
|---|---|---|
| 4282096 | Major depressive disorder, single episode | MDD-qualifying diagnosis |
| 4282316 | Major depressive disorder, recurrent episode | MDD-qualifying diagnosis |
| 436665 | Bipolar disorder | BD-qualifying diagnosis; disqualifying for MDD |
| 443237 | Manic episode | BD-qualifying diagnosis; disqualifying for MDD |
| 435783 | Schizophrenia | Disqualifying (both clinical cohorts) |
| 4286201 | Schizoaffective disorder | Disqualifying (both clinical cohorts) |
| 438409 | Attention deficit hyperactivity disorder (ADHD) | Disqualifying (both clinical cohorts) |
| 439776 | Autism spectrum disorder (ASD) | Disqualifying (both clinical cohorts) |
| 440069 | Drug dependence | Disqualifying (both clinical cohorts); also used to define NCC exclusion |
| 4182210 | Dementia | Disqualifying (both clinical cohorts); also used to define NCC exclusion |
| 443432 | Cognitive impairment | Disqualifying (both clinical cohorts); also used to define NCC exclusion |
| 4209423 | Nicotine dependence | Explicit exception: not disqualifying |
| 437264 | Tobacco dependence | Explicit exception: not disqualifying |
| 432586 | Mental disorder (ancestor concept) | Used to define the Healthy Control exclusion criterion (any descendant diagnosis excludes a participant from NCC) |

Table S3: **Sample retention cascades for the MDD, BD, and NCC cohorts**. Percentages indicate data retention at each sequential filtering step relative to the preceding row. Notably, the single largest reduction in sample size occurs when requiring baseline Fitbit data prior to the EtM battery. This drop reflects the limited overlap between the *All of Us* subpopulation that completed the cognitive tasks and the subpopulation using wearable devices. Due to the small final sample size realised for the BD cohort, we did not pursue the relationship between wearable phenotypes and cognitive tasks in this group. For the (well-powered) NCC cohort, this evaluation was conducted only exploratorily as a sensitivity analysis since our focus is on cognitive functioning in mood disorders. The colour code shows the sample size available to study cognitive deviation relative to NCC and relationship between cognitive measures and wearable phenotypes.

(a) MDD cohort.

| | GradCPT | Flanker | DD | EmoRecog |
|---|---|---|---|---|
| Completed EtM task | 7,641 | 7,109 | 7,625 | 9,020 |
| No mood episode within 90 days of test | 5,618 (73.5%) | 5,220 (73.4%) | 5,598 (73.4%) | 6,624 (73.4%) |
| Test session passed QC | 5,546 (98.7%) | 5,151 (98.7%) | 5,475 (97.8%) | 6,622 (100.0%) |
| Covariates available (age, sex, education, device) | 5,472 (98.7%) | 5,087 (98.8%) | 5,411 (98.8%) | 6,536 (98.7%) |
| $\geq$90 days of Fitbit data preceding test | 1,250 (22.8%) | 1,182 (23.2%) | 1,249 (23.1%) | 1,528 (23.4%) |
| Fitbit window free of mood episodes | 1,138 (91.0%) | 1,094 (92.5%) | 1,146 (91.8%) | 1,392 (91.1%) |
| Covariates available (BMI, CCI, latitude) | 1,080 (94.9%) | 1,036 (94.7%) | 1,084 (94.6%) | 1,320 (94.8%) |
| $\geq$30 valid days/nights in 90-day window | 544 (50.4%) | 516 (49.8%) | 538 (49.6%) | 641 (48.6%) |

(b) BD cohort.

| | GradCPT | Flanker | DD | EmoRecog |
|---|---|---|---|---|
| Completed EtM task | 1,036 | 955 | 1,076 | 1,280 |
| No mood episode within 90 days of test | 610 (58.9%) | 568 (59.5%) | 644 (59.9%) | 763 (59.6%) |
| Test session passed QC | 601 (98.5%) | 558 (98.2%) | 625 (97.0%) | 762 (99.9%) |
| Covariates available (age, sex, education, device) | 584 (97.2%) | 545 (97.7%) | 608 (97.3%) | 739 (97.0%) |
| $\geq$90 days of Fitbit data preceding test | 170 (29.1%) | 161 (29.5%) | 174 (28.6%) | 203 (27.5%) |
| Fitbit window free of mood episodes | 149 (87.6%) | 148 (91.9%) | 158 (90.8%) | 186 (91.6%) |
| Covariates available (BMI, CCI, latitude) | 136 (91.3%) | 134 (90.5%) | 143 (90.5%) | 170 (91.4%) |
| $\geq$30 valid days/nights in 90-day window | 75 (55.1%) | 73 (54.5%) | 74 (51.7%) | 87 (51.2%) |

(c) NCC cohort.

| | GradCPT | Flanker | DD | EmoRecog |
|---|---|---|---|---|
| Completed EtM task | 61,030 | 61,030 | 61,030 | 61,030 |
| Test session passed QC | 44,111 (72.3%) | 41,152 (67.4%) | 43,339 (71.0%) | 52,247 (85.6%) |
| Covariates available (age, sex, education, device) | 43,515 (98.6%) | 40,589 (98.6%) | 42,734 (98.6%) | 51,491 (98.6%) |
| $\geq$90 days of Fitbit data preceding test | 8,233 (18.9%) | 7,758 (19.1%) | 8,145 (19.1%) | 9,650 (18.7%) |
| Covariates available (BMI, CCI, latitude) | 7,716 (93.7%) | 7,297 (94.1%) | 7,651 (93.9%) | 9,028 (93.6%) |
| $\geq$30 valid days/nights in 90-day window | 4,276 (55.4%) | 4,033 (55.3%) | 4,210 (55.0%) | 4,932 (54.6%) |

Table S4: **Demographic characteristics of the healthy control (NCC) sample**. Reassuringly, the age, sex, education, and device-type-of-administration distributions in the NCC sample span the ranges observed in the MDD and BD cohorts, supporting the use of the NCC-derived normative model to compute demographically matched deviation scores ($D$) for clinical participants without requiring substantial extrapolation beyond the range of the data on which the model was fit.

(a) Sample for the NCC normative model (Section 2.5.1).

| Variable | GradCPT ($N$ = 43,515) | Flanker ($N$ = 40,589) | DD ($N$ = 42,734) | EmoRecog ($N$ = 51,491) |
|---|---|---|---|---|
| Age (years) | 59.6 (±15.9) | 59.4 (±15.9) | 59.2 (±16.1) | 58.5 (±16.3) |
| Female sex | 28,543 (65.6%) | 26,353 (64.9%) | 28,156 (65.9%) | 34,335 (66.7%) |
| Education (0–7) | 6.0 (±1.0) | 6.0 (±1.0) | 6.0 (±1.0) | 6.0 (±1.0) |
| Touchscreen | 25,979 (59.7%) | 24,531 (60.4%) | 25,723 (60.2%) | 31,431 (61.0%) |
| Ethnicity: | | | | |
| White | 34,499 (79.3%) | 32,092 (79.1%) | 33,779 (79.0%) | 40,449 (78.6%) |
| Black | 2,123 (4.9%) | 1,986 (4.9%) | 2,196 (5.1%) | 2,676 (5.2%) |
| Asian | 1,480 (3.4%) | 1,429 (3.5%) | 1,436 (3.4%) | 1,788 (3.5%) |
| Task score | 2.7 (±0.8) | 0.4 (±0.2) | −5.5 (±1.8) | 0.9 (±0.1) |

(b) Sample for the NCC sensitivity analysis on the wearable-phenotype–cognition association (Section 2.7).

| Variable | GradCPT ($N$ = 4,276) | Flanker ($N$ = 4,033) | DD ($N$ = 4,210) | EmoRecog ($N$ = 4,932) |
|---|---|---|---|---|
| Age (years) | 60.7 (±15.1) | 60.5 (±15.2) | 60.4 (±15.3) | 59.8 (±15.4) |
| Female sex | 2,756 (64.5%) | 2,564 (63.6%) | 2,686 (63.8%) | 3,205 (65.0%) |
| Education (0–7) | 6.1 (±1.0) | 6.1 (±0.9) | 6.1 (±1.0) | 6.1 (±0.9) |
| Touchscreen | 2,673 (62.5%) | 2,566 (63.6%) | 2,665 (63.3%) | 3,134 (63.5%) |
| BMI (kg/$m^2$) | 28.3 (±6.6) | 28.3 (±6.6) | 28.4 (±6.6) | 28.4 (±6.7) |
| CCI | 1.0 (±2.2) | 1.1 (±2.3) | 1.0 (±2.2) | 1.0 (±2.3) |
| Season: | | | | |
| Spring | 5.6% | 5.9% | 6.3% | 6.3% |
| Summer | 18.6% | 18.6% | 19.1% | 17.2% |
| Autumn | 62.8% | 62.2% | 61.4% | 63.0% |
| Winter | 13.1% | 13.3% | 13.3% | 13.5% |
| Ethnicity: | | | | |
| White | 3,277 (76.6%) | 3,072 (76.2%) | 3,220 (76.5%) | 3,743 (75.9%) |
| Black | 271 (6.3%) | 250 (6.2%) | 268 (6.4%) | 315 (6.4%) |
| Asian | 234 (5.5%) | 220 (5.5%) | 240 (5.7%) | 292 (5.9%) |
| Wearable Features: | | | | |
| Steps (mean/day) | 7,597.8 (3,831.3) | 7,623.4 (3,828.1) | 7,646.5 (3,896.9) | 7,616.6 (3,844.5) |
| WASO (min) | 56.7 (13.2) | 56.4 (13.1) | 56.4 (13.3) | 56.4 (13.0) |
| TST (min) | 392.8 (52.1) | 392.5 (52.6) | 392.5 (52.9) | 392.5 (52.8) |
| STV (h) | 1.1 (0.5) | 1.1 (0.5) | 1.1 (0.5) | 1.1 (0.5) |
| Task score | 2.7 (±0.8) | 0.4 (±0.2) | −5.6 (±1.7) | 0.1 (±0.1) |

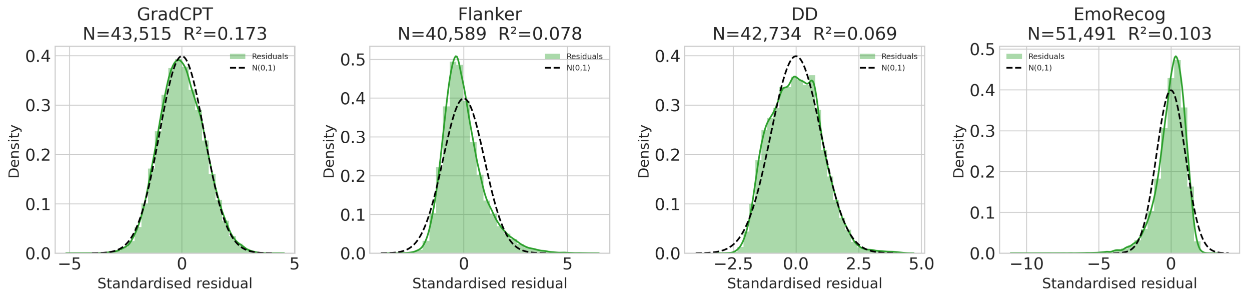


Figure S1: **Standardised residuals per task across healthy controls.** For each task in the *EtM* battery, we regressed raw task scores in the NCC cohort on the demographic and administration-mode covariates we sought to control for: age, sex, education, and touchscreen device use (Section 2.5.1). The resulting residuals were then standardised to zero mean and unit variance ($N$ and model $R^2$ shown above each panel). This same regression equation, fitted strictly on NCC data, was applied to MDD and BD participants to compute each individual's normative deviation score $D$, representing the number of NCC residual standard deviations by which observed performance differs from the demographically matched expectation (Section 2.5.1). Residual distributions approximate $\mathcal{N}(0, 1)$ (dashed reference curve) most closely for GradCPT, whereas Flanker and EmoRecog show comparatively higher departure from normality. This departure does not affect the validity of downstream analyses. Group-mean and contrast tests in Section 2.5.1 employ heteroscedasticity-robust (HC3) standard errors rather than relying on a normal-residual assumption, and with $N$ 4,000 NCC and $N$ 5,00 MDD participants per task, the sampling distributions of these coefficients and of the Pearson correlations in Section 2.5.2 are governed by the Central Limit Theorem rather than by the shape of the underlying residual distribution. Normal-theory percentile interpretations of individual $D$ values should nonetheless be applied cautiously for Flanker and EmoRecog specifically.

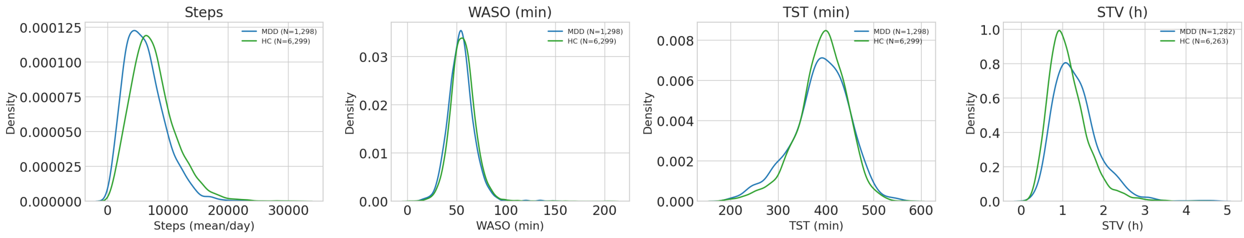


Figure S2: **Wearable phenotypes distribution across MDD and NCC**. We present the distribution of wearable phenotypes for the MDD and NCC groups. While a formal comparative analysis between MDD and NCC is beyond the scope of this paper, we note that the two groups display visibly distinct distributions in Steps, STV, and TST, whereas no substantial differences emerge for WASO. The direction of these observational differences is consistent with previous literature.

### Minimum Detectable Effect Size

Table S5: Minimum detectable effect sizes (MDES) are computed at both a nominal significance level ($\alpha = 0.05$) and a Bonferroni-corrected level ($\alpha = 0.05/28$), giving optimistic and conservative bounds respectively; the true detection threshold under the Benjamini-Hochberg procedure used for our primary analyses lies between these two bounds. Section 2.5.1 tests group means and their contrast, expressed as $\Delta_{\text{Glass}}$ (consistent with $D$ already being expressed in NCC-residual-SD units; see Section 2.5.1). Section 2.5.2 tests a bivariate association (Pearson $r$ and $R^2 = r^2$). In (b) and (c), only nominal-threshold MDES are shown, as these analyses are not part of the main 28-test pool. Table cells are colour-coded inversely to the MDES: darker shading indicates smaller MDES, i.e., greater power to detect subtle effects.

(a) Primary analyses (main 28-test pool).

| | | | $\Delta_{\text{Glass}}$ | | Pearson $r$ | |
|---|---|---|---|---|---|---|
| Task | Group / Phenotype | $N$ | nom. | Bonf. | nom. | Bonf. |
| *Cognition Difference Relative to NCC (Section 2.5.1) — effect size: $\Delta_{\text{Glass}}$* | | | | | | |
| GradCPT | MDD | 5,472 | 0.038 | 0.054 | — | — |
| | BD | 584 | 0.116 | 0.165 | — | — |
| | Contrast | – | 0.122 | 0.173 | — | — |
| Flanker | MDD | 5,087 | 0.039 | 0.056 | — | — |
| | BD | 545 | 0.120 | 0.171 | — | — |
| | Contrast | – | 0.126 | 0.179 | — | — |
| DD | MDD | 5,411 | 0.038 | 0.054 | — | — |
| | BD | 608 | 0.114 | 0.161 | — | — |
| | Contrast | – | 0.120 | 0.170 | — | — |
| EmoRecog | MDD | 6,536 | 0.035 | 0.049 | — | — |
| | BD | 739 | 0.103 | 0.146 | — | — |
| | Contrast | – | 0.109 | 0.154 | — | — |
| *Association between Cognition and Wearable Phenotypes (Section 2.5.2) — effect size: Pearson $r$* | | | | | | |
| GradCPT | Steps / WASO / TST | 544 | — | — | 0.120 | 0.169 |
| | STV | 540 | — | — | 0.120 | 0.169 |
| Flanker | Steps / WASO / TST | 516 | — | — | 0.123 | 0.173 |
| | STV | 512 | — | — | 0.124 | 0.174 |
| DD | Steps / WASO / TST | 538 | — | — | 0.121 | 0.170 |
| | STV | 535 | — | — | 0.121 | 0.170 |
| EmoRecog | Steps / WASO / TST | 641 | — | — | 0.110 | 0.156 |
| | STV | 636 | — | — | 0.111 | 0.156 |

(b) Associations between wearable phenotypes and cognitive performance in NCC (Section 2.7).

| Task | Phenotype | $N$ | $r$ (nom.) |
|---|---|---|---|
| GradCPT | Steps / WASO / TST / STV | 4,276 | 0.043 |
| Flanker | Steps / WASO / TST / STV | 4,033 | 0.044 |
| DD | Steps / WASO / TST / STV | 4,210 | 0.043 |
| EmoRecog | Steps / WASO / TST / STV | 4,932 | 0.040 |

(c) Associations between wearable phenotypes and cognitive tasks (Section 2.7) in BD. This is powered to detect effect sizes of a magnitude unrealistic in this setting(Table S5a).

| Task | Phenotype | $N$ | $r$ (nom.) |
|---|---|---|---|
| GradCPT | Steps / WASO / TST | 75 | 0.319 |
| | STV | 75 | 0.319 |
| Flanker | Steps / WASO / TST | 73 | 0.323 |
| | STV | 73 | 0.323 |
| DD | Steps / WASO / TST | 74 | 0.321 |
| | STV | 74 | 0.321 |
| EmoRecog | Steps / WASO / TST | 87 | 0.297 |
| | STV | 87 | 0.297 |

### Sensitivity Analysis

All results in the sensitivity analysis are reported at nominal (uncorrected) $p$-values and should be interpreted accordingly.

#### Differences between MDD population included in and excluded from Fitbit analysis

We tested for differences in cognitive deviation ($D$) and covariates between the MDD population included in the wearable phenotype analysis and the MDD population excluded from it, separately for each cognitive task. Only education showed a nominal difference between included and excluded participants, for the Delay Discounting task ($p = 0.0462$; shaded). Overall, these results reassuringly show that they are no systematic difference across the two populations.

Table S6: **MDD population included in vs. MDD population excluded from wearable phenotype analysis**.

| Variable | Included | Excluded | Test | Statistic | Effect size | $p$ |
|---|---|---|---|---|---|---|
| **GradCPT** | | | | | | |
| | $N = 544$ | $N = 4{,}928$ | | | | |
| $D$ | $-0.067 \pm 0.994$ | $-0.082 \pm 0.987$ | Welch $t$ | 0.336 | +0.015 | 0.7367 |
| Age at EtM | $58.250 \pm 14.883$ | $58.344 \pm 15.087$ | Welch $t$ | −0.140 | −0.006 | 0.8891 |
| Sex (% female) | 24.8 | 24.0 | $\chi^2$ | 0.134 | +0.005 | 0.7138 |
| Education | $5.897 \pm 1.027$ | $5.840 \pm 1.003$ | Welch $t$ | 1.231 | +0.057 | 0.2188 |
| BMI | $31.053 \pm 7.870$ | $30.707 \pm 7.685$ | Welch $t$ | 0.974 | +0.045 | 0.3305 |
| CCI | $3.178 \pm 3.776$ | $3.179 \pm 3.734$ | Welch $t$ | −0.002 | −0.000 | 0.9988 |
| Touchscreen (%) | 66.4 | 63.9 | $\chi^2$ | 1.205 | +0.015 | 0.2724 |
| **Flanker** | | | | | | |
| | $N = 516$ | $N = 4{,}571$ | | | | |
| $D$ | $0.032 \pm 1.034$ | $-0.032 \pm 0.984$ | Welch $t$ | 1.331 | +0.064 | 0.1838 |
| Age at EtM | $58.207 \pm 14.967$ | $58.228 \pm 15.079$ | Welch $t$ | −0.030 | −0.001 | 0.9764 |
| Sex (% female) | 25.0 | 24.6 | $\chi^2$ | 0.026 | +0.002 | 0.8712 |
| Education | $5.874 \pm 1.009$ | $5.817 \pm 1.012$ | Welch $t$ | 1.209 | +0.056 | 0.2270 |
| BMI | $31.010 \pm 7.883$ | $30.683 \pm 7.628$ | Welch $t$ | 0.897 | +0.043 | 0.3703 |
| CCI | $3.324 \pm 3.923$ | $3.146 \pm 3.688$ | Welch $t$ | 0.979 | +0.048 | 0.3280 |
| Touchscreen (%) | 66.7 | 65.7 | $\chi^2$ | 0.153 | +0.005 | 0.6956 |
| **DD** | | | | | | |
| | $N = 538$ | $N = 4{,}873$ | | | | |
| $D$ | $0.035 \pm 1.005$ | $0.098 \pm 1.015$ | Welch $t$ | −1.375 | −0.062 | 0.1695 |
| Age at EtM | $57.757 \pm 14.984$ | $57.909 \pm 15.288$ | Welch $t$ | −0.223 | −0.010 | 0.8235 |
| Sex (% female) | 25.3 | 23.9 | $\chi^2$ | 0.399 | +0.009 | 0.5275 |
| Education | $5.900 \pm 1.014$ | $5.808 \pm 1.021$ | Welch $t$ | 1.998 | +0.090 | 0.0462 |
| BMI | $30.924 \pm 7.932$ | $30.760 \pm 7.668$ | Welch $t$ | 0.458 | +0.021 | 0.6474 |
| CCI | $3.243 \pm 3.841$ | $3.157 \pm 3.718$ | Welch $t$ | 0.499 | +0.023 | 0.6183 |
| Touchscreen (%) | 66.5 | 64.6 | $\chi^2$ | 0.734 | +0.012 | 0.3916 |
| **EmoRecog** | | | | | | |
| | $N = 641$ | $N = 5{,}895$ | | | | |
| $D$ | $0.048 \pm 0.904$ | $0.033 \pm 0.980$ | Welch $t$ | 0.391 | +0.015 | 0.6960 |
| Age at EtM | $57.546 \pm 14.650$ | $57.405 \pm 15.357$ | Welch $t$ | 0.231 | +0.009 | 0.8176 |
| Sex (% female) | 25.3 | 24.0 | $\chi^2$ | 0.468 | +0.008 | 0.4938 |
| Education | $5.872 \pm 1.027$ | $5.818 \pm 1.018$ | Welch $t$ | 1.267 | +0.053 | 0.2055 |
| BMI | $31.099 \pm 7.918$ | $30.705 \pm 7.732$ | Welch $t$ | 1.196 | +0.051 | 0.2319 |
| CCI | $3.240 \pm 3.837$ | $3.142 \pm 3.743$ | Welch $t$ | 0.619 | +0.026 | 0.5359 |
| Touchscreen (%) | 68.0 | 65.5 | $\chi^2$ | 1.523 | +0.015 | 0.2171 |

#### Association between cognitive tasks and wearable phenotypes in NCC

Owing to its much larger sample size, the NCC sample was better powered to detect small effects than the MDD sample (Table S5b). Associations were in the expected direction: higher physical activity (Steps), longer TST, and less day-to-day variability in sleep timing (STV) were associated with better sustained attention (GradCPT) and lower impulsivity (DD); STV was similarly associated with better emotion recognition accuracy (EmoRecog). Flanker showed no association with any wearable phenotype. These associations, while statistically significant at nominal (uncorrected) $p$-values, were small in magnitude, with correlation coefficients ($r$) ranging from $-0.108$ (STV, GradCPT) to $+0.064$ (TST, GradCPT), corresponding to at most $\sim$1.2% of variance explained ($R^2$). Thus, statistical significance here should not be read as evidence of a clinically meaningful relationship.

Table S7: **Association between Wearable Phenotypes and Cognitive Tasks in NCC**.

| **Task** | **Fitbit Phenotype** | **Group** | **NCC ($N$)** | $\beta$ | **ES ($r$)** | $R^2$ | $p_{\textbf{raw}}$ |
|---|---|---|---|---|---|---|---|
| **GradCPT** | Steps | NCC | 4,276 | +0.059 | +0.060 | 0.0035 | 0.0001178 |
| | WASO | NCC | 4,276 | -0.004 | -0.004 | 0.0000 | 0.797 |
| | TST | NCC | 4,276 | +0.063 | +0.064 | 0.0041 | 3.83e-05 |
| | STV | NCC | 4,256 | -0.107 | -0.108 | 0.0117 | 4.137e-13 |
| **Flanker** | Steps | NCC | 4,033 | +0.008 | +0.008 | 0.0001 | 0.616 |
| | WASO | NCC | 4,033 | +0.019 | +0.019 | 0.0004 | 0.2039 |
| | TST | NCC | 4,033 | +0.013 | +0.014 | 0.0002 | 0.3822 |
| | STV | NCC | 4,013 | -0.016 | -0.016 | 0.0003 | 0.3014 |
| **DD** | Steps | NCC | 4,210 | -0.036 | -0.037 | 0.0014 | 0.01747 |
| | WASO | NCC | 4,210 | -0.001 | -0.001 | 0.0000 | 0.955 |
| | TST | NCC | 4,210 | -0.047 | -0.048 | 0.0023 | 0.001827 |
| | STV | NCC | 4,190 | +0.052 | +0.053 | 0.0028 | 0.0008126 |
| **EmoRecog** | Steps | NCC | 4,932 | +0.010 | +0.011 | 0.0001 | 0.4371 |
| | WASO | NCC | 4,932 | -0.010 | -0.011 | 0.0001 | 0.438 |
| | TST | NCC | 4,932 | +0.026 | +0.028 | 0.0008 | 0.06213 |
| | STV | NCC | 4,909 | -0.045 | -0.048 | 0.0023 | 0.001758 |

#### Assessing presence of a U-shaped relationship between TST and cognitive measures

A common assumption in sleep research is that cognitive performance is best at some intermediate TST, with both too little and too much sleep associated with worse performance – a "U-shaped" relationship. Our primary analyses (Section 2.5.2) tested only for a straight-line (linear) association between rest-activity phenotypes and cognitive deviation. As a sensitivity check, we additionally tested TST specifically for this U-shaped pattern, separately in the MDD and NCC samples.

We did this by adding a curvature term to the standard model and asking three questions of the result, all of which must hold for the data to genuinely support a U-shape rather than a simple straight-line trend or an artefact of the model: **A) Is there real curvature?** We tested whether the relationship between TST and cognitive deviation bends significantly, rather than following a straight line. **B) Does the curve's turning point fall within the range of TSTs we actually observed?** A model can mathematically produce a curve, but if the point where it turns over lies far outside the TSTs seen in our participants, this is not evidence of a genuine 'sweet 'spot, it is an extrapolation beyond the data. **c) Are both extremes (very short and very long sleep) actually associated with worse cognitive performance than the middle?** A curve can bend without both ends being worse than the middle (e.g., cognition could plateau at one extreme rather than decline).

Across both samples and all four cognitive tasks, we found consistent evidence for a TST "sweet spot" in two instances, both in the well-powered NCC sample: sustained attention (GradCPT) and emotion recognition (EmoRecog). In both cases, cognitive performance was highest at an intermediate, commonly observed TST and declined at both shorter and longer durations. No evidence for a U-shape was found for Flanker or Delay Discounting in NCC, nor for any task in the MDD sample. Taken together, this suggests that a U-shaped TST–cognition relationship, where present, is

Table S8: **Test for a U-shaped relationship between TST and cognitive deviation, by task and sample** "Curvature present" reflects the significance ($p < 0.05$, uncorrected) of the quadratic term. "Turning point observed" indicates whether the curve's vertex falls within the 5th–95th percentile range of observed TST values. "Both extremes worse" indicates whether predicted cognitive deviation at both the 5th and 95th percentile of TST was worse than at the turning point, in the direction consistent with impairment for that task. A U-shape is considered supported only when all three criteria are met.

| Task | Sample | $N$ | Curvature present? | Turning pt. observed? | Both ext. worse? | U-shape supported |
|---|---|---|---|---|---|---|
| GradCPT | MDD | 544 | No ($p = 0.759$) | No | No | No |
| GradCPT | NCC | 4,276 | Yes ($p < 0.001$) | Yes | Yes | Yes |
| Flanker | MDD | 516 | No ($p = 0.678$) | Yes | No | No |
| Flanker | NCC | 4,033 | No ($p = 0.088$) | Yes | No | No |
| DD | MDD | 538 | No ($p = 0.386$) | Yes | Yes | No |
| DD | NCC | 4,210 | No ($p = 0.127$) | Yes | Yes | No |
| EmoRecog | MDD | 641 | No ($p = 0.501$) | No | No | No |
| EmoRecog | NCC | 4,932 | Yes ($p = 0.025$) | Yes | Yes | Yes |

detectable only in the better-powered NCC arm within the range of TSTs captured by our wearable data, and that the MDD sample's null on this check does not rule out a similarly-sized non-linearity that it lacks the power to resolve.
